# Unique Metabolic Profiles Associate with Gestational Diabetes and Ethnicity in Low and High-Risk Women Living in the UK

**DOI:** 10.1101/2022.04.11.22273658

**Authors:** Harriett Fuller, Mark Iles, J. Bernadette Moore, Michael A. Zulyniak

## Abstract

**Background:** Gestational Diabetes Mellitus (GDM) is the most common global pregnancy complication; however, prevalence varies substantially between ethnicities with South Asians (SA) experiencing up to 3-times the risk of the disease compared to white Europeans (WEs). Factors driving this discrepancy are unclear, although the metabolome is of great interest as GDM is known to be characterised by metabolic dysregulation.

**Objective:** This primary aim was to characterise and compare the metabolic profiles of GDM in SA and WE women (at < 28 weeks’ gestation) from the Born in Bradford (BIB) prospective birth cohort in the UK.

**Methods:** 146 fasting serum metabolites, from 2668 pregnant WE and 2671 pregnant South Asian (SA) women (average BMI 26.2 kg/m^2^, average age 27.3 years) were analysed using partial least squares discriminatory analyses to characterise GDM status. Linear associations between metabolite values and post-oral glucose tolerance test measures of dysglycemia (fasting glucose and 2-hour post glucose) were also examined.

**Results:** Seven metabolites associated with GDM status in both ethnicities (variable importance in projection (VIP) ≥1), while 6 additional metabolites associated with GDM only in WE women. Unique metabolic profiles were observed in healthy weight women who later developed GDM, with distinct metabolite patterns identified by ethnicity and BMI status. Of the metabolite values analysed in relation to dysglycemia, lactate, histidine, apolipoprotein A1, HDL cholesterol, HDL2 cholesterol associated with decreased glucose concentration, while DHA and the diameter of very low-density lipoprotein particles (nm) associated with increased glucose concertation in WE women; while in SAs albumin alone associated with decreased glucose concentration.

**Conclusions:** This study shows that the metabolic risk profile for GDM differs between WE and SA women enrolled in BiB the UK. This suggests that aetiology of the disease differs between ethnic groups and that ethnic-appropriate prevention strategies may be beneficial.

## Introduction

During pregnancy, there is a natural increase in catabolism to ensure sufficient energy for the foetus (1, 2). This increase is governed by maternal hormones, beginning as a mild change in insulin sensitivity and progressing through hyperinsulinemia to controlled insulin resistance by the third trimester (2-5). For most pregnancies, these changes are safe and controlled, with insulin sensitivity returning to a healthy state following pregnancy. However, for approximately one in seven pregnancies, insulin resistance exceeds normal “healthy” levels and enters a diabetic state, putting the mother and her growing offspring in danger of short- and long-term health risks (6, 7). This pregnancy-induced state of diabetes, gestational diabetes mellitus (GDM), is a major global health concern with varying prevalence between populations.

In Middle Eastern, North Africa, and South Asian countries, GDM prevalence can exceed 20% of pregnancies, whereas in European countries prevalence of GDM is more commonly around 5% (5). Numerous lifestyle, biological, and genetic factors are thought to contribute to this disparity of risk (5, 8). Despite the numerous factors, diet is the mainstay of most prevention and treatment strategies because of its demonstrated efficacy for managing glucose concentrations (9-11). Nonetheless, we and others have demonstrated that the effects of dietary prevention strategies on maternal and offspring health are not generalisable across populations or ethnic groups, with dietary patterns demonstrating varied effects between ethnic groups in relation to both GDM prevention and birth weight (12-15). These data suggest that the metabolism and pathology of GDM may differ across populations, where some ethnic groups have unique metabolic profiles that make them more susceptible to GDM (4, 5, 16-18). Specifically, elevated concentrations of alanine, numerous fatty acids (e.g., myristic acid, palmitic acid, palmitoleic acid) and lower amounts of glutamate, proline, and phospholipids in blood have been identified as predictors of GDM risk in early pregnancy (i.e., before 16 weeks) (4), with recent evidence demonstrating significant differences in the abundance of these metabolites between ethnic groups (19). Notably, evidence from Born in Bradford (BiB), a prospective multi-ethnic pregnancy and birth cohort, has demonstrated the need for potentially modified GDM assessment criteria for South Asian (SA) women because of increased risks of delivery complication and newborn macrosomia at significantly lower glucose thresholds, compared to white European (WE) women (20). Indeed, currently the UK’s National Health System (NHS), routinely screens all women of South Asian ancestry for GDM while only high-risk WE women are screened (21).

As a consequence of this, the Diabetic Pregnancy Study Group called for increased research into the role of the metabolome on GDM in 2018 (22). To date, however, the metabolic drivers of GDM remain unclear with numerous discrepancies within the field, likely due to small, heterogenous cohorts of heterogenous cohorts of varying populations, cultures, and ancestral groups (23). Indeed, only one study has conducted an analysis of individual metabolites and GDM in an ethnic-specific fashion (1). This work investigated univariate associations between numerous metabolites in WE (n = 4072) and SA (n = 4702) women and demonstrated that concentrations of lipoproteins and cholesterols are typically higher in WE women and are stronger predictors of GDM (ie. have a higher VIP score), compared to SA women. However, metabolite profiles are heterogenous mixtures of metabolites, many of which are strongly correlated and may depend on other metabolites to exhibit an effect. In light of this, multivariate approaches that assess all variables simultaneously along with their inter-variable correlations (24) can be used to identify (i) patterns of uncorrelated metabolites that associate with GDM risk, and (ii) cardinal metabolites that independently associate with GDM risk. Therefore, this study aims to build upon existing work by applying multivariate statistical techniques within an ethnically diverse population to (i) determine underlining metabolite patterns that correlate with GDM, (ii) identify ethnic-specific metabolic drivers of GDM risk.

## Materials and Methods

### i. Population Characteristics

The BiB cohort was established to examine determinants of health from pregnancy and childhood into adulthood in an ethnically diverse region in the north of England (25). Between 2007 and 2010, BiB recruited 12,453 women (26-28 weeks’ gestation, mean maternal age 27.8), collecting baseline data on 13,776 pregnancies and 13,858 births, with 45% of the cohort of SA origin (25, 26). BiB aimed to recruit all mothers giving birth at the Bradford Royal Infirmary, the largest hospital within Bradford. Bradford is a northern English city with high levels of deprivation and a large SA population, the majority of which have Pakistani ancestry. All women were invited to partake in an oral glucose tolerance test (OGTT) for GDM diagnosis at approximately 26-28 weeks during their standard antennal care. Almost all UK citizens utilize the NHS for antenatal care.

Of these, 11,480 women provided blood samples for metabolite analyses during the same visit as their OGTT. Written consent was gained from all participants and ethical approval was granted by the Bradford Research Ethnics Committee (ref07/H1302/112)(25).

### ii. Blood Metabolite Analysis

Full details of venous blood sample collection, preparation, metabolite quantification and validation have previously been described in detail (1). In brief, fasted blood samples were taken at the Bradford Royal Infirmary by trained phlebotomists, processed within 2.5 hours and stored at -80°C in the absence of freeze-thaw cycles. (27) Samples were processed using a high-throughput automated NMR platform and have previously been validated (Nightingale Health©; Helsinki, Finland). Metabolite values expressed as a percentage or ratio were excluded to minimize redundancy, resulting in a panel of 146 metabolite values expressed in absolute quantitative measures. This panel comprised measures of 97 lipoproteins, 9 amino acids, 2 apolipoproteins, 9 cholesterols, 8 fatty acids, 8 glycerides and phospholipids, 4 glycolysis-related metabolites, 2 ketone bodies, 3 measures of fluid balance and inflammation, and 3 measures of the mean lipoprotein particle diameter (**Supplemental S1**).

### iii. Participant Selection

Of the 11,480 blood samples analyzed for metabolites, 54 samples were excluded because they failed one of five Nightingale© quality control measures (low glucose, high lactate, high pyruvate, low protein concentration and plasma samples). Of the 11,426 available samples, ∼3% of mothers were missing ≥1 metabolite values. The structure of missing metabolite data was assessed via the visualization and imputation of missing values (VIM) package within R (28) and multiple correspondence analysis (MCA). There was no evidence that the missing metabolite data occurred in a non-random pattern. It was therefore deemed appropriate to impute missing values. Optimized multiple imputation with iterative principal component analysis (PCA; 100 simulations, K-fold cross validation) based upon the minimization of mean square error of prediction (MSEP) was performed using the missMDA package (29). A sensitivity analysis was performed to test the effect of mothers with higher rates of missingness (≥3% missing metabolite values) on imputation. No detectable difference in imputation quality was noticed. As such, the metabolite data of all available 11,426 maternal samples were included for imputation.

Imputed metabolite data were combined with descriptive BiB reported characteristics, including participant’s ethnicity, age moved to UK (if born abroad) GDM status, gestational age at sample collection (obtained from obstetric records), history of diabetes, age, BMI, smoking status, parity and whether they were carrying a singleton/multiple pregnancy. Length of residence was calculated by subtracting the age the mother moved to the UK from maternal age. When an individual was born within the UK, length of residency was taken to be mothers age.

All women were recruited prior to their scheduled GDM assessment (mean gestational age 26.1 weeks), and prior to the 28th week of pregnancy. GDM was diagnosed using a modified version of the World Health Organization criteria (1, 25). Using this criteria, a women was diagnosed with GDM if either their fasting glucose concentration exceed ≥ 6.1 mmol/L or if 2-hour post-load glucose concentrations was ≥ 7.8 mmol/L following a 75g oral glucose tolerance test (OGTT). The OGTT was completed in the morning following an overnight fast and involved the consumption of a standard solution over a 5-minute period containing the equivalent of 75g of anhydrous glucose (30). Following a GDM diagnosis all SA and WE mothers receive the same standardised care following a GDM diagnosis. Initially GDM management involves referral to a dietitian and the management of glucose concentration through diet and increased exercise. If unsuccessful, managed by metformin or insulin injections will be prescribed. Women with GDM will also be offered additional antenatal appointments to monitor the health of both mother and baby throughout the pregnancy. Irrespective of GDM status, basic nutritional counselling is offered to all mothers as part of standard antenatal classes offered throughout pregnancy by the UK National Health Services (NHS) (31).

Ethnicity was self-reported. If ethnicity was not collected, details were obtained from primary care records along with information on parity and the number of registered births. Maternal age was recorded at pregnancy booking (ie., the first routine antenatal visit) and BMI was calculated using height measured at recruitment and maternal weight recorded at the first antenatal visit. When examined as a categorical variable, ethnic specific cut-offs were used to classify mothers into BMI groups (underweight: ≤ 18.5 kg/m^2^ in WE and SAs, normal/healthy weight: 18.6-25 kg/m^2^ in WEs or 18.6-22.9 kg/m^2^ in SAs, overweight: 25-29.9 kg/m^2^ for WE or 23-27.4 kg/m^2^ for SA women; obese: > 30kg/m^2^ for WE or >27.5kg/m^2^ for SA women) (32). When analysed as a binary variable, women were grouped as having a ‘healthy’ or ‘high’ BMI if they were above/below the BMI cut-off for overweight status utilizing these ethnic-specific cut offs. Smoking status was self-reported at baseline and during pregnancy. Recruitment and the baseline assessment of covariates was the same in both ethnic groups. Summary statistics for each variable were presented as a mean and standard error (SE). Difference in baseline characteristics were calculated between women with and without GDM for continuous variables via a Mann-Whitney (MW) test, while differences for categorical variables were tested using Pearson’s Chi-squared test.

Participants whose samples were collected after GDM diagnosis (28^th^ week or later) were excluded from the analysis as well as mothers with a history of diabetes. Individuals who reported being of a South Asian origin other than Pakistani (SA) were also excluded, due to the small sample size (therefore limited power) of other South Asian ancestry groups. In total, 5,339 participants, 2,671 SAs (all of Pakistani descent) and 2,688 White European (WE) women, were retained for analysis. **(Figure 1)**

**Figure 1:**
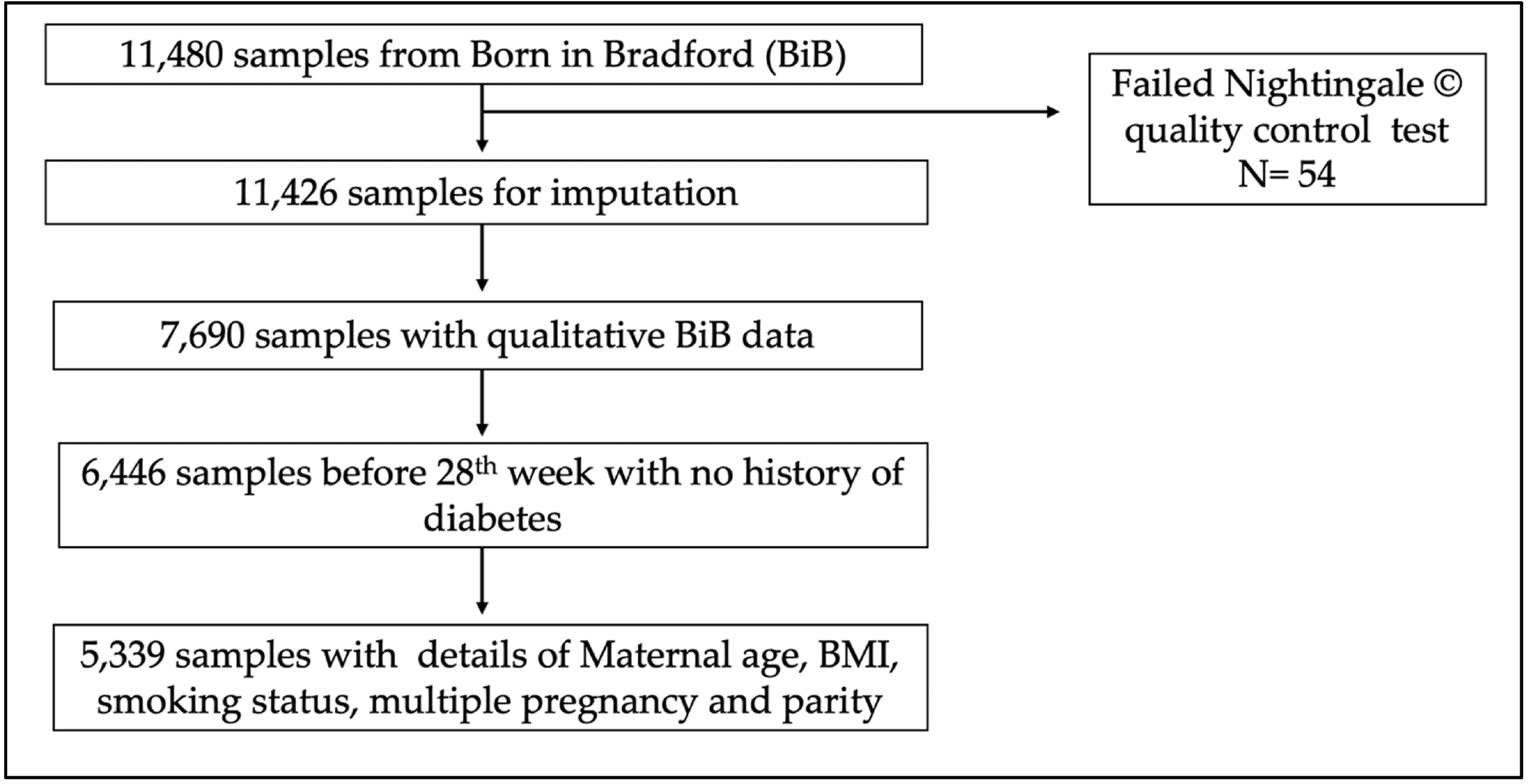
Flow chart of study participants from the Born in Bradford (BiB) cohort included within this study.

Ethnicity was self-reported and the homogeneity of the WE group has been confirmed in previous genetic analyses within BIB (33). 93.2% of the included WEs were born in the British Isles (i.e., the UK, Republic of Ireland, Channel Islands or Isle of Man), with the majority in England (91.4%). Of these women, 95.5% reported that both of their parents were also born in the British Isles. Within the group of WE women not born in the British Isles, 3.7% were born in Eastern Europe (Czech Republic, Poland, Slovakia) with the remaining proportion reporting ‘other’ or ‘unknown’. Within the SA population, 43.7% were born within the UK. Of the SA women born in the UK, 93% reported that their mother was born in Pakistan (87.4%) or India (5.6%) and 95% reported that their father was born in Pakistani (88.6%) or India (6.7%). A small proportion did not know their mother’s (1.4%) or father’s (1.3%) place of birth. Of the women born outside of the UK, the average age of immigration to the UK was 18.8 yrs (IQR 18 – 23).

### iv. Metabolite Discriminatory Analysis

Partial least squares discriminatory analysis (PLSDA) is a supervised dimensionality reduction technique that uses all included variables to discriminate group data based upon predefined outcome groups. Included variables are then ranked by the degree to which they explain the variance between groups (i.e., GDM vs non-GDM). These are known as variable importance in projection (VIPs), where VIPs ≥1 denote a variable with good discriminatory quality and predictive ability (34, 35).

PLSDA allowed an overall assessment of the predictive capacity of metabolites for GDM, in models with and without known GDM risk factors (i.e., BMI, maternal age, parity, multiple pregnancy, and smoking status), with ethnicity added to visually assess its effect on the model. Following this, both sets of PLSDA models were performed within each ethnic group. To assess bi-directionality, models predicting ethnicity were also executed within the overall population and GDM cases/ non-cases separately using the same criteria as above.

The optimum number of components to include within the model was selected based upon the component’s ability to significantly predict group membership within the training (pR^2^Y ≤0.05) and validation (pQ^2^Y≤ 0.05) datasets (7-fold cross validation, ‘nipals’ algorithm). When multiple components were significantly predictive, the predictive component that best discriminated between groups (i.e. maximization of outcome variance explained, R^2^Y) with the minimal error (root mean squared error of estimation (RMSEE)) was selected. Data were pareto scaled and mean-centered prior to analysis. External validity was assessed via 7-fold cross validation. PLSDA models were performed via the ‘ropls’ package within R (36). When the size of the outcome groups differed by ≥ 1% the larger group was randomly sampled (n=20) to minimize error. VIPs were mean averaged and SEs calculated across all significant iterations (pR^2^Y ≤0.05, pQ^2^Y≤0.05) for each metabolite following the removal of outlier VIPs, defined as 1.5 x interquartile range of VIP values. Differences in the distribution of VIP values between both ethnicities and case-status were assessed for significant iterations via a MW test; this was possible because all comparisons were tested against the same panel of metabolite measures. To assess the impact of smoking on PLSDA results, PLSDA models predicting smoking in the overall study population were also performed.

### iii. Post-Hoc multivariate analyses

BMI is a suspected mediator along the casual pathway that links metabolism and GDM, was a significant driver of GDM within SA women and WE women. To explore this, the ethnic-specific impact of BMI on the metabolome and subsequent GDM diagnoses was investigated using sparse PLSDA (sPLSDA). sPLSDA is a supervised multivariate technique with the ability to predict group membership in multiclass problems (i.e., stratification by ethnicity, bodyweight, and GDM status) by simultaneously performing and balancing variable selection with group classification (37). Women were classified as ‘healthy or ‘overweight’ based upon ethnic-specific cut-offs (BMI ≥ 25kg/m^2^ for WE women and BMI≥ 23kg/m^2^ for SA women), which is the same approach used by the NHS (38). The analyses focussed on low-risk WE (n= 872) and low-risk SA women (n= 864) — i.e., only women (i) in their first pregnancy, (ii) that did not smoke during pregnancy, and (iii) were < 35 years of age were included. This was done to prevent these covariates from overpowering the models, and allowing the contributing roles of BMI on GDM to be more clearly appreciated within and between each ethnic group.

Metabolites selected by sPLSDA in each comparison were fed into PLSDA models (20 iterations) alongside highly correlated metabolites (Pearsons correlation coefficient ≥0.9) in order determine metabolie values contributing to the separation of the outcome groups whilst balancing dimensionality reduction and group discrimination. PLSDA models were adjusted for maternal age (continuous), BMI (continous), smoking status, partiy and multiple pregnancy such as before. Differences in the distributions of metabolites within each group were also compared by a MW test.

### iii. Linear regression analyses for identified metabolite associations

Linear regression models investigating the relationship between post-oral OGTT measures (fasting glucose and 2-hour post-OGTT) were performed on all metabolite values identified as important (VIP≥1) in characterizing GDM status. Normality of glucose measures and metabolite values were assessed using histograms and Q-Q plots. Most metabolites (136/146) required normalization. Normality was most often achieved by log transformation (59 metabolite values); however, in some cases square-root and normal score transformation (NST) were implemented via the ‘rcompanion’ package(39). All glucose measures were log normalized. Known GDM risk factors of maternal age (years), gestational age (days), parity, and smoking status during pregnancy (yes/no), were initially including in the models. When significant associations were observed between metabolite values and glucose in this exploratory analysis (P<0.05), BMI was added to the models (initially as a continuous and then as a binary variable utilizing ethnic-specific BMI cut-offs for overweight status) to assess the role of early pregnancy BMI as a mediator of metabolite-dysglycemia associations. Within SAs, a final additional adjustment of length of residency within the UK was made to account for any effects of acculturation.

## Results

### i. Population Characteristics

The mean age of participant was 26.7 years and had a mean BMI of 26 kg/m^2^. WE women were significantly older and had higher BMIs compared to SA women (**Table 1**). Parity was significantly higher in SA women compared to WE women (P<0.001) and parity was only significantly higher in GDM cases compared to non-cases within SA women (**Table 1**). Smoking during pregnancy was significantly more common in WE women compared to SA women (25% vs 3%; P<0.001). No difference in proportion of singleton pregnancies (>97%) was observed between WE women and SA women. Alcohol intake was not assessed because it was reported by only 1% of SA women. The mean time of sample collection was 187 gestational days.

**Table 1:** Population Characteristics at < 28 weeks’ gestation (mean gestational age 26.7 weeks) from the Born in Bradford (BiB) cohort.^2^.

|  | Overall | White European |  |  |  | South Asian |  |  |  | White European vs South Asian P-values |  |  |
| --- | --- | --- | --- | --- | --- | --- | --- | --- | --- | --- | --- | --- |
|  | Total (n=5339) | Total (n=2668) | GDM (n=128) | Non-GDM (n=2540) | P-value | Total (n=2,671) | GDM (n=286) | Non-GDM (n=2385) | P-value | Overall (n=5339) | GDM (n=414) | Non-GDM (n=4925) |
| **Age (years)** | 27.3 (0.08) | 26.7 (0.1) | 30 (0.5) | 26.5 (0.1) | < 0.001 | 25.7 (0.1) | 30.6 (0.3) | 27.6 (0.1) | < 0.001 | < 0.001 | 0.29 | < 0.001 |
| **Mothers weight at baseline assessment (kg)** | 68.8 (0.2) | 72.0 (0.3) | 76.8 (1.6) | 71.8 (0.3) | 0.002 | 65.5 (0.3) | 72.3 (0.9) | 64.7 (0.3) | < 0.001 | < 0.001 | 0.03 | < 0.001 |
| **Mothers' height (cm)** | 162 (0.09) | 164.3 (0.1) | 163.8 (0.5) | 164.3 (0.1) | 0.47 | 159.7 (0.1) | 158.1 (0.3) | 159.9 (0.1) | < 0.001 | < 0.001 | < 0.001 | < 0.001 |
| **BMI<sup>1</sup>** |  |  |  |  |  |  |  |  |  |  |  |  |
| *mean (kg/m<sup>2</sup>)* | 26.2 (0.08) | 26.7 (0.1) | 28.5 (0.5) | 26.6 (0.1) | < 0.001 | 25.7 (0.1) | 28.9 (0.4) | 25.3 (0.1) | < 0.001 | < 0.001 | 0.49 | < 0.001 |
| *underweight or normal* | 2198 (41.2) | 1254 (47) | 47 (36.7%) | 1207 (47.5%) | 0.02 | 944 (35.3) | 46 (16.1) | 898 (37.7) | < 0.001 | < 0.001 | < 0.001 | < 0.001 |
| *overweight or obese* | 3141 (58.8) | 1414 (53) | 81 (63.3%) | 1333 (52.5%) |  | 1727 (64.7) | 240 (83.9) | 1487 (62.3) |  |  |  |  |
| **Parity** |  |  |  |  | 0.47 |  |  |  | < 0.001 | < 0.001 | < 0.001 | < 0.001 |
| 0 | 2311 (43.2) | 1394 (52.2) | 73 (57) | 1321 (52) |  | 917 (34.3) | 82 (28.7) | 835 (36.5) |  |  |  |  |
| 1 | 1508 (28.2) | 813 (30) | 39 (30.5) | 774 (30.5) |  | 695 (26) | 49 (17.1) | 646 (28.3) |  |  |  |  |
| 2 | 813 (15.2) | 293 (11) | 11 (8.6) | 282 (11.1) |  | 520 (19.5) | 57 (19.9) | 463 (20.3) |  |  |  |  |
| ≥ 3 | 707 (13.2) | 168 (6.3) | 5 (3.9) | 163 (10.4) |  | 539 (20.2) | 98 (34.3) | 441 (14.9) |  |  |  |  |
| **Singleton pregnancy (%)** | 5274 (98.8) | 2634 (98.7) | 123 (96.1) | 2511 (98.9) | 0.01 | 2640 (98.8) | 280 (97.9) | 2360 (99) | 0.12 | 0.70 | 0.29 | 0.75 |
| **Smoked during pregnancy (%)** | 958 (17.9) | 870 (32.6) | 25 (19.5) | 845 (33.3) | 0.001 | 88 (3.3) | 12 (4.2) | 76 (3.2) | 0.37 | < 0.001 | < 0.001 | < 0.001 |

<sup>1</sup> Summary table of population characteristics, expressed as a mean (SE) for continuous variables and counts (%) for categorical variables. Differences between women with and without GDM for continuous variables were tested using a Mann-Whitney test, while differences for categorical variables were tested using Pearson's Chi-squared test.

### ii. Primary Analysis

#### Metabolite Characterisation of GDM

In the 1st model, an overall analysis of the full cohort (i.e., both ethnic groups), PLSDA explained 21.7% of the variation between the GDM and non-GDM groups and confirmed maternal age and BMI as primary risk factors for GDM risk followed by parity, smoking status, and having a non-singleton pregnancy as the primary drivers of GDM (**Table 2**). In the full model, 7 metabolite values reported VIPs ≥ 1, including 4 fatty acid metabolite measures (total fatty acids, 18:2 linoleic acid, total MUFA and total SFA), and one glycolysis related metabolite (lactate) (**Figure 2**). Modelled independently, the PLSDA with only covariates explained 12.4% of the variation in GDM status and significantly predicted GDM status, whereas the model with only metabolites explained 13.5% of the variance in GDM but was non-significant. The 2^nd^ model, which included ethnicity as a covariate, accounted for 26.6% of the variation between the GDM and non-GDM groups. The same 6 metabolites we reported as predictors of GDM with an additional cholesterol metabolite measure (total esterified cholesterol). Notably, model 2 confirmed ethnicity (SA vs WE) as a major risk factor for GDM, after age and BMI. Modelled independently, ‘ethnicity’ and other covariates explained 15.2% of the variance in GDM status; therefore, the addition of metabolites into the model increased the amount of variance explained by over 11%.

**Table 2:** Key metabolite measures (VIP ≥1) that discriminate women diagnosed as GDM from non-GDM women in Partial Least Squares Discriminatory Analysis (PLSDA).^1^.

| Variable | Model 1 | Model 2 |
| --- | --- | --- |
| Age | 6.4 (0.03) | 5.9 (0.03) |
| BMI | 5.4 (0.04) | 5.1 (0.02) |
| Ethnicity | - | 2.9 (0.02) |
| Parity | 2.4 (0.01) | 2.3 (0.01) |
| Smoking Status | 1.9 (0.02) | 1.7 (0.01) |
| Multiple Pregnancy | 1.5 (0.01) | 1.3 (0.009) |
| Lactate | 1.5 (0.01) | 1.2 (0.008) |
| VLDL_D | 1.3 (0.01) | 1.3 (0.01) |
| Total Fatty Acids | 1.2 (0.01) | 1.5 (0.01) |
| Total MUFA | 1.2 (0.001) | 1.2 (0.008) |
| 18:2 Linoleic Acid | 1.1 (0.01) | 1.1 (0.004) |
| Total SFA | 1.1 (0.01) | 1.2 (0.007) |
| Esterified Cholesterol | - | 1.0 (0.008) |

<sup>1</sup> Variables of Importance in Projection (VIPs) scores [mean and (standard error)] across 20 PLSDA iterations discriminating between GDM (n=414) and non-GDM women (n=414). Model 1: Includes all metabolite measures plus BMI (continuous), maternal age (years), smoking status, parity and multiple pregnancy status. Model 2: Model 1 + ethnicity. GDM, Gestational Diabetes Mellitus; MUFA, total monounsaturated fatty acids; SFA, total saturated fatty acids; VLDL\_D, mean diameter of very-low density lipoproteins.

**Figure 2:**
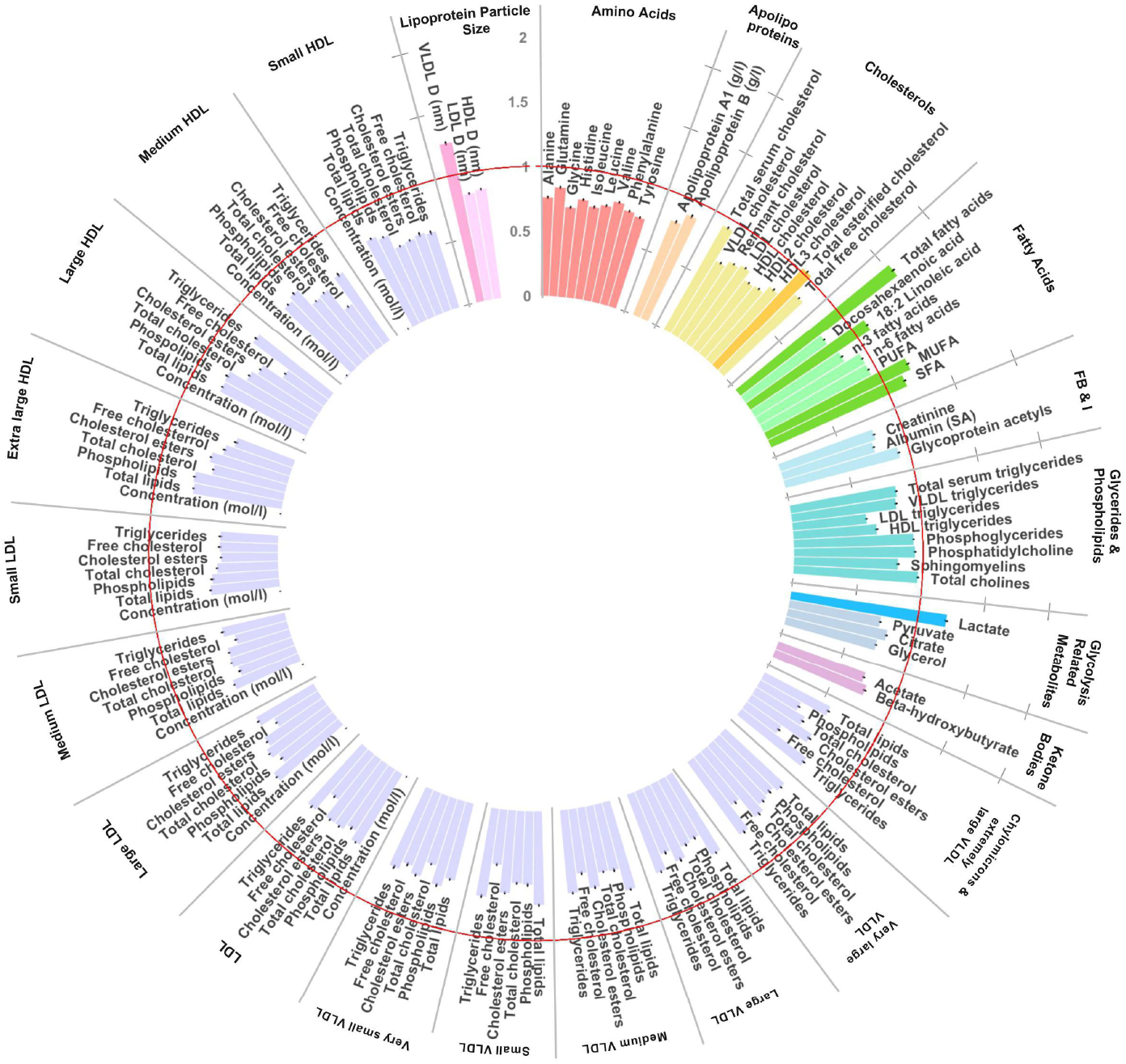
Circular bar plot identifying key metabolites (VIP ≥ 1) that distinguished 414 women with gestational diabetes mellitus (GDM) from 414 women without GDM. Mean average VIP scores across 20 PLSDA model iterations (n_cases_ = 414). Bars represent standard errors (SEs). The PLSDA included maternal age (years), BMI (continuous), smoking status, parity, and multiple pregnancy status, and ethnicity. Red line denotes VIP cut-off of 1. Units mmol/L unless stated. GRM, Glycolysis Related Metabolites; LPS, Lipoprotein Particle Size; MUFA, total monounsaturated fatty acids; SFA, total saturated fatty acids; VLDL_D, mean diameter of very-low density lipoproteins.

#### Ethnically-stratified analysis of metabolites characterising GDM

In an ethnically stratified analysis (20 iterations), models only including metabolites accounted for a median average of 6.5% of the variation in GDM status in SA women and 5.8% of the variation in WE women in optimised models (ie. minimisation RMSEE and maximisation of R^2^Y) although no model comprising metabolites alone was signficant. Conversely, models only including established clinical risk factors (age, BMI, parity, smoking status and multiple pregnancy) were significantly predictive (p-value R^2^ < 0.05, Q^2^<0.05) of GDM status and explained 13.3% of the variation in SAs 12.8% of the variation in WEs. The addition of metabolites to these covariate models also resulted in the significant prediction of GDM. These models resulted in 26% of the variance in GDM status in WE women and 20% of the variance in SA women being accounted for, an increase of 13.6% and 6.8% when compared to covariate models in WE and SA respectively. Following adjustments for maternal age, parity, BMI, and smoking status, GDM could be predicted within both ethnicites. Maternal age, parity and BMI were predictors of GDM in both ethnicities (VIP≥1), with BMI the most important predictor of GDM in SA women, while in WE women maternal age was most important predictor (**Supplementary table 1**). Smoking was a predictor of GDM only in WE women. After adjustment for confounders, 7 metabolite variables characterised GDM status (VIP≥1) in both ethnicities (total fatty acids, total MUFA, total SFA, linoleic acid, glycoprotein acetyls, lactate, and diameter of VLDL) (**Figure 3, Supplementary table 2**). Of these metabolites, the VIPs of three (lactate, glycoprotein acetyls, and linoleic acid) characterised GDM status comparatively well between ethnicites (VIP≥1; MW P>0.05), whereas four metabolite measures (total fatty acids, total MUFA, total SFA, and VLDL_D) characterised GDM in both ethnic groups, but were significantly stronger markers of GDM in WE women (VIP≥1; MW P<0.05 between ethnicites). Additionally, alanine, glutamine, total cholesterol, total n-6 PUFA, total PUFA, and citrate were markers (VIP≥1) of GDM status in WE women only. No markers of GDM were specific to SA women. On average, the optimised models explained 26% of the variance of GDM in WE women, and 20% of the variance in SA, women (**Supplementary table 3**).

**Figure 3:**
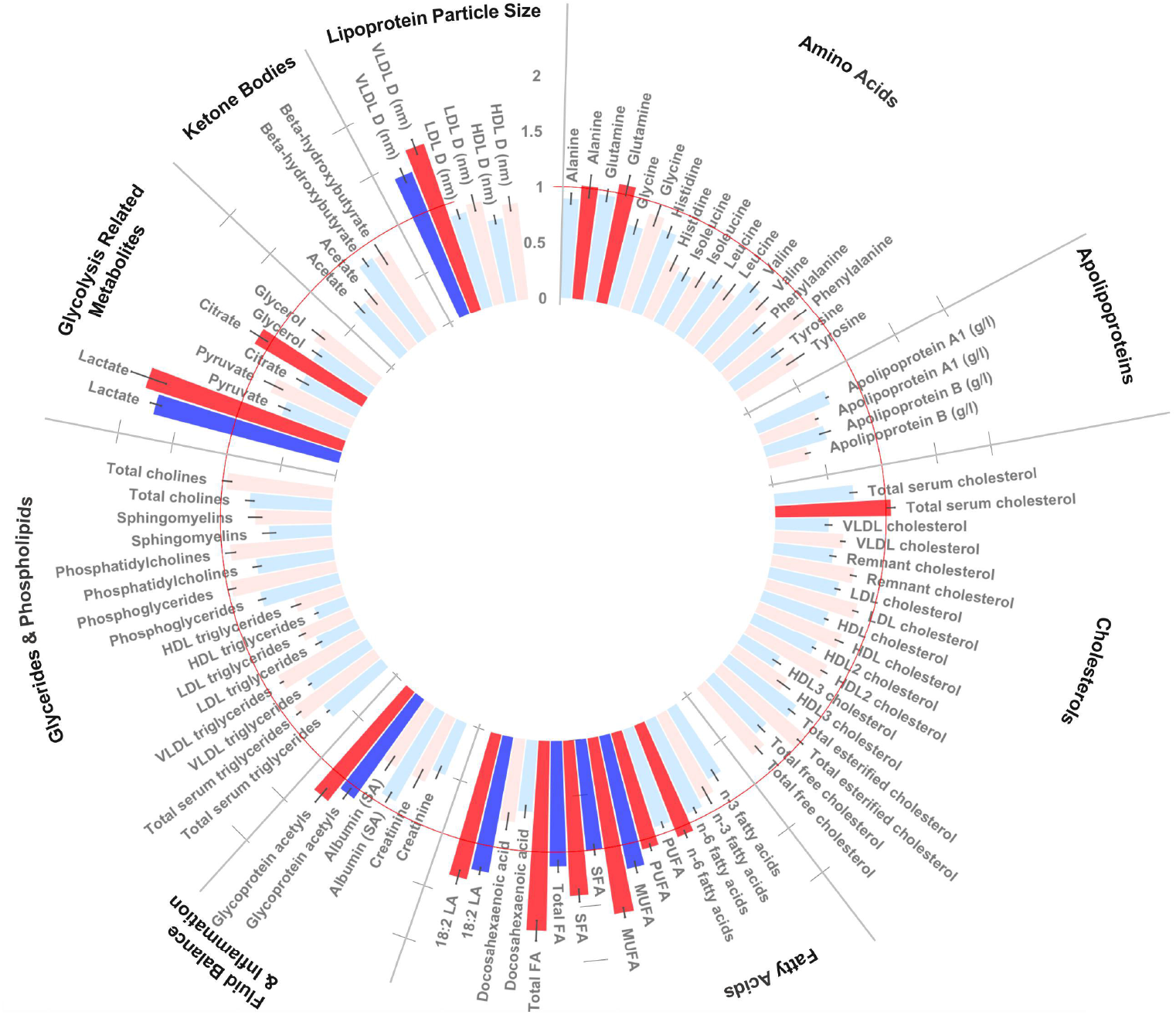
Circular bar plot of ethnically stratified analyses identifying key metabolites (VIP ≥ 1) that distinguished GDM women from non GDM women in South Asians (n_cases_=286) 2and white Europeans (n_cases_=128). Mean average VIP scores across 20 PLSDA model iterations (n_casesSA_ = 286, n_casesWE_=128). Bars represent standard errors (SEs). The PLSDA was run separately for SA (blue) and WE (red) women and included maternal age (years), BMI (continuous), smoking status, parity, and multiple pregnancy status. Red circular line denotes VIP cut-off of 1. No lipoproteins demonstrated a VIP >1 and were not included in the figure to preserve space. Units mmol/L unless stated. 18.2 LA, 18.2 Linoleic acid; GRM, Glycolysis Related Metabolites; LPS, Lipoprotein Particle Size; MUFA, total monounsaturated fatty acids; SFA, total saturated fatty acids; Tot FA, Total fatty acids; VLDL_D, mean diameter of very-low density lipoproteins.

#### Metabolites characterised by ethnicity

To explore underlying metabolic profiles within each ethnic group, we identified metabolites that most strongly distinguished WE women and SA women. In a PLSDA including known GDM risk factors as covariates (maternal age, smoking status, parity, BMI, and GDM status), 12 metabolic measures VIP≥1 in statistically significant models (models p-values R^2^ > 0.05 and Q^2^>0.05) and therefore were believed to have characterised ethnicity in GDM and non-GDM women: total fatty acids, serum cholesterol, SFA, MUFA, FAw6, esterified cholesterol, LA, LDL cholesterol, remnant cholesterol, phosphatidylcholine and total cholesterol. (**Supplementary table 4**).

Additionally, ethnicity was characterised by 6 metabolites values exclusivley in women diagnosed with GDM [i.e alanine, total fatty acids, linoleic acid (LA), glycoprotein acetyls, lactate and diameter of VLDL] whilst 5 metabolites values were exclusive in those not diagnosed with GDM [i.e., apolipoprotein A1, remnant choelsterol, docosahexanoic acid (DHA), and phosphatidylcholine]. An additional 9 metabolite values [ie., total serum cholesterol, LDL cholesterol, total esterfied cholesterol, n-3 fatty acids, PUFA, MUFA, SFA, phosphatidylcholine and total cholines] were predictive of ethnicity in both GDM cases and non-cases (**Supplementary figure 1**).

### iv. Post-Hoc Analyses

#### Characterisation of GDM in Low-Risk Women

BMI was classifed as an important variable (VIP ≥ 1) in the overall analysis and in both ethnic subgroup analyses. However, a greater mean VIP (± SE) was observed in SA women compared to WE women (VIP_SA_ = 7.06 ± 0.22 vs. VIP_WE_ = 4.33 ± 0.22; P<0.001) (**Supplementary table 1**) indicating that BMI may be a more important predictor of GDM status within WEs. Indeed, healthy weight SA women who developed GDM (SA_Healthy-GDM_) presented the most distinct metabolic profile (Receiver Operator Curve; ROC = 0.783), but were most similar to healthy WE women who developed GDM (WE_Healthy-GDM_; ROC = 0.691) (**Supplementary figure 2**). The reason for this shared and distinct pattern of metabolites in ‘healthy’ weight women who developed GDM is unclear and many hypotheses are possible. One hypothesis may be that the pattern is an artifact of their fetal programming. Adult offspring from GDM pregnancies are at increased risk of dysglycemia, diabetes, and GDM that has been attributed to metabolic dysregulation, and early dysglycemia that progresses in later life (40-43).

Future work in established cohorts that investigate trans-generational pregnancy risks (such as Born in Bradford, Generation R, and Nutrigen) are integral to unravel the source of this unique metabolic profile which distinguishes healthy weight GDM cases of SA ancestry from non-cases, overweight SA cases, and WE cases (25, 44, 45). Due to the higher proportion of underweight mothers of SA ancestry, a sensitivity analysis was performed where underweight mothers were removed (n_removed_ = 93, BMI≤18.5 kg/m^2^) to determine if their profiles were unique. No difference in the outcome was observed following the removal of these individuals.

Metabolites selected by sPLSDA in each comparison were fed into PLSDA models (20 iterations) alongside highly correlated metabolites (Pearsons correlation coefficient ≥0.9) to identify key metabolic drivers of this separation (**Supplementary figure 3**). Alanine, glutamine, and glycerol were important to distinguish healthy weight SA women who developed GDM (SAC-N) from all others, whereas fatty acids were important to distinguish SAC-N from other GDM cases. Interestingly, in healthy women, aromatic and branched chain amino acids distinguished GDM and non-GDM women between (but not within) ethnic groups. Glycerol distributions were significantly different in all comparisons (MW <0.05).

#### Characterisation of GDM in low-risk women by BMI and ethnicity

Orthogonal partial least squares discriminant analysis (oPLSDA) is a supervised multivariate technique that separates variation within each predictor variables based upon its linear (correlated) and orthogonal (uncorrelated) association with the outcome variable (46, 47). This can provide better separation along fewer components when a large proportion of variance within the dataset does not directly correlate with the outcome variable. Furthermore, through the creation of Shared and Unique Structure (SUS) plots it is possible to determine shared and unique factors separating the main group of interest (healthy weight SA cases, SAC-N) with the two most relevant biological comparisons (healthy weight SA non-cases, SANC-N and healthy weight WE cases, WEC-N).

No significant separation of the SAC-N vs SANC-N, SAC-N vs SAC-H and SAC-N vs WEC-N groups were identified via SUS plots with oPLSDA. Following the inclusion of BMI and age within the models the SAC-N group was found to separate from all other groups (**Supplementary figure 4**). BMI was the only variable found to be responsible for this separation with a high magnitude and reliability. Pyruvate, L-HDL and XL-HDL had a small impact on the separation of the SAC-N group but with a low reliability, as shown within SUS-plots.

#### Association between important metabolites and gestational dysglycemia

Overall, 8 of 146 metabolite measures were associated with fasting glucose or 2-hour post glucose (**Table 3**), all of which were identified as GDM predictors via PLSDA or sPLSDA. The analysis in WE women demonstrated the greatest number of associations between metabolite and glucose measures. Six metabolites positively associated fasting glucose concentration (albumin, lactate, histidine, apolipoprotein A1, HDLC and HDL2C), while one negatively associated with fasting glucose (mean density of LDL) (**Supplementary table 5, Supplementary table 6)**. Only DHA associated with 2-hr post OGTT in WEs, where a 1 mmol/L increase in DHA associated with a 0.20 mmol/L increase in 2-hour post glucose. In the analysis of SA women, only albumin was associated with dysglycemia, where higher albumin associated with lower concentration of fasting glucose and 2-hr post OGTT. In an additional analysis, length of residency within the UK was added to the fully-adjusted model to evaluate the role of UK acculturation as a modifier of the association between albumin and postprandial glucose measures. In both models significant associations were identified with albumin (P-value_fasting_ =0.031, β= -0.79, SE=0.37, P-value_2-hour_ =0.028 β=-1.75, SE=0.80). Length of residency was not found to be a significant variable in the model, but the magnitude of associations decreased slightly following its inclusion (**Supplementary table 5, Supplementary table 6**). In the ethnic-combined analysis, associations between albumin, lactate, and mean diameter of LDL with fasting glucose retained significance. Adjusting for BMI as a continuous or binary variable had no impact on the associations.

**Table 3:**
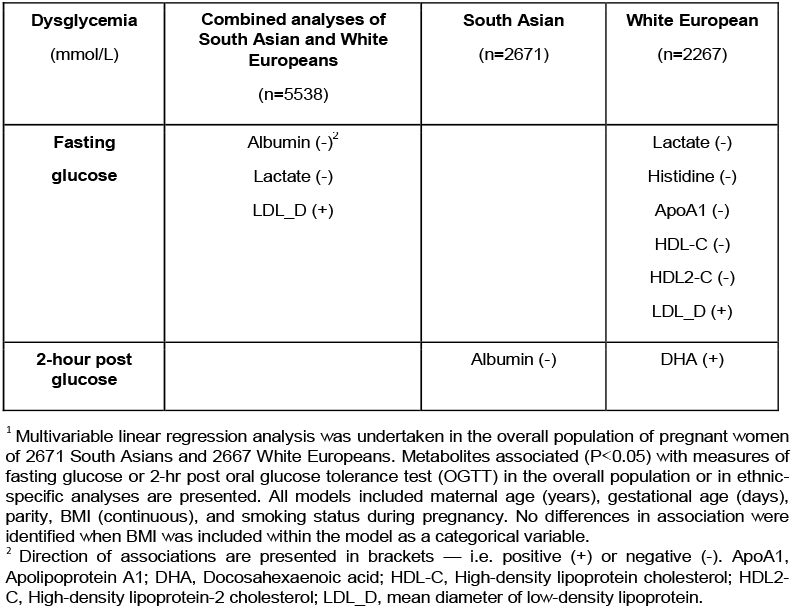
Metabolite measures associated with dysglycemia in South Asian and white European pregnant women before 28 weeks’ gestation (mean gestational age 26.7 weeks)^1^.

| **Dysglycemia**<br>(mmol/L) | **Combined analyses of**<br>**South Asian and White**<br>**Europeans**<br>(n=5538) | **South Asian**<br>(n=2671) | **White European**<br>(n=2267) |
| --- | --- | --- | --- |
| **Fasting**<br>**glucose** | Albumin (-)<sup>2</sup><br>Lactate (-)<br>LDL_D (+) |  | Lactate (-)<br>Histidine (-)<br>ApoA1 (-)<br>HDL-C (-)<br>HDL2-C (-)<br>LDL_D (+) |
| **2-hour post**<br>**glucose** |  | Albumin (-) | DHA (+) |

<sup>1</sup> Multivariable linear regression analysis was undertaken in the overall population of pregnant women of 2671 South Asians and 2667 White Europeans. Metabolites associated ( $P < 0.05$ ) with measures of fasting glucose or 2-hr post oral glucose tolerance test (OGTT) in the overall population or in ethnic-specific analyses are presented. All models included maternal age (years), gestational age (days), parity, BMI (continuous), and smoking status during pregnancy. No differences in association were identified when BMI was included within the model as a categorical variable.
<sup>2</sup> Direction of associations are presented in brackets — i.e. positive (+) or negative (-). ApoA1, Apolipoprotein A1; DHA, Docosahexaenoic acid; HDL-C, High-density lipoprotein cholesterol; HDL2-C, High-density lipoprotein-2 cholesterol; LDL\_D, mean diameter of low-density lipoprotein.

## Discussion

Using a prospective birth cohort with an equal proportion of WE and SA women, we identified 7 metabolite measures that characterized GDM in both WE and SA women — 4 of which were more predictive in WE women. These results agree with the Omega cohort (78.5% non-Hispanic white; nested case-control; 46 cases, 47 controls) that highlighted a distinct metabolic profile at 16-weeks’ gestation (comprised of fatty acids, sugars, alcohols, amino acids and organic acids), associated with future GDM diagnosis (48). Although the metabolite patterns identified by the Omega study were not predictive, our predictive multivariate analysis (and a previous univariate analyses) (1) found similar associations between GDM and many of these metabolites (i.e., amino acids, glycolysis related metabolites, and fatty acids), and offers further evidence of ethnic-specific associations.

Given the overall elevated risk of GDM observed in SA women compared to WE women, even at a healthy BMI (i.e., OR≈3) (49), and the role of ethnicity in predicting GDM, in the present study, we sought to characterize distinct metabolic profiles of SA and WE women. Of the 146 metabolite values tested, 7 were important for stratifying GDM and non-GDM women in the overall population (lactate, mean density of VLDL particles, total fatty acids, total MUFAs, 18:2 linoleic acid, total SFA and esterified cholesterol). Following stratification by ethnicity, alanine, glutamine, total serum cholesterol, n-6 fatty acids, PUFAs, and citrate distinguished GDM and non-GDM in WE women whereas no metabolite values were predictive solely within SA women.

Although no metabolite value identified solely within WE women was associated with post-OGTT measures of glucose in *post-hoc* analyses, our evidence agrees with previous work from (i) a small case-control study (26 T2Ds vs 7 controls) that reported alanine, glutamine, and citrate to characterize GDM and controls, with citrate being a key marker of diabetics with underlying complications (e.g., CVD) (50), and (ii) a cohort study of 431 pregnant Chinese women (12-16 weeks’ gestation), where alanine and glutamine were associated with GDM (51). Biologically, alanine, glutamine, and citrate are connected and could moderate dysglycemia through their interaction with the tricarboxylic acid cycle (TCA) to promote the formation of TCA intermediates, fatty acid synthesis, and modulate glucagon and insulin secretion (52, 53). Taken together, it may be that alanine and glutamine are more robust markers of dysglycemia, whereas citrate is a marker of metabolic or physiologic stress in diabetic individuals — such as pregnancy. The role of total cholesterol is uncertain as it is not convincingly associated with dysglycemia (a meta-analysis of 73 observational studies found no association)(54), suggesting that associations between total cholesterol and GDM are complex and/or subject to confounding.

In the ethnic subgroup analyses, fatty acids were identified as the most important family (ie. VIP ≥ 1) to characterize GDM status. In WE women and SA women respectively, 75% and 50% of the fatty acids included within the metabolite panel were considered ‘important’ to characterize GDM within WE women. Furthermore, in SA women fatty acids constituted more than half of all metabolites with a VIP ≥ 1.

This reflects earlier work by Taylor *et al*. [1], which identified some evidence of ethnic specific associations between fatty acids and GDM, and agrees with molecular analyses that demonstrate that fatty acids alter insulin resistance and insulin secretion during pregnancy (55, 56).

Furthermore, fatty acids (total MUFAs, total n-3 PUFAs, total n-6 PUFAs, total PUFAs, and DHA) were identified as key metabolic factors to distinguish healthy-weight SA and WE women who developed GDM. Interestingly, we highlighted associations between n-6 PUFA and total PUFAs with GDM that were specific to WE women. Given the equal sample sizes between groups, and that fatty acids were important to characterize ethnicity, it is suggestive of ethnic differences in PUFA metabolism (57-59) and a role in ethnic-associated GDM risk (57, 60, 61). Indeed, n6 PUFA-derived eicosanoids show discriminatory qualities between type-2 diabetics and controls with good accuracy (R^2^X = 0.824, R^2^Y = 0.995, Q^2^ = 0.779) and were identified as proposed mediators of dysglycemia within a Chinese population (62). Longitudinal analyses to evaluate the association between changes in PUFA and eicosanoids concentrations on dysglycemia during pregnancy are required to better understand this association.

The association between VLDL diameter and dysglycemia is supported by a recent hypothesis linking insulin resistance, triglyceride synthesis, and increased VLDL diameter (63, 64). Although we cannot disregard that VLDLs are sensitive to level of fasting (65) (*as our participants were subjected to prior to blood collection*), evidence also suggests that ethnic-specific genetic variants associate with ethnic-specific differences in VLDL diameter (66). Although there has been less work on the possible association between glycoprotein acetyls (a marker of systemic inflammation) and GDM, and future work is required in this area

Lactate was one of the strongest predictors of GDM within both groups, in agreement with evidence from a case-control study in China (n=12 GDM; n=10 controls) (67) and pathway analyses that propose lactate as a regulator of insulin resistance and a marker metabolic syndrome severity (68, 69). *Post-hoc* analysis demonstrated no association between glycoprotein acetyls and glucose concerntrations, while lactate and mean diameter of VLDL were associated with fasting glucose in WE women but not SA women. The multi-ethnic HAPO cohort demonstrated a similar ethnic-specific association between lactate and fasting glucose within individuals of Northern European ancestry but not minority ethnic groups (48, 70, 71).

Of the numerous fatty acid measures that were associated with GDM, only DHA was associated with a post-OGTT measures of glucose and only in WE women. Overall, DHA is considered a protective metabolite against insulin resistance (e.g., HOMA-IR); however, recent evidence suggests high heterogeneity (56, 72, 73). As we did, researchers investigating the Camden pregnancy cohort (n=1,368) reported a significant positive linear association between DHA and HOMA-IR (0.303 ± 0.152 per unit DHA %; P<0.05) (56), while conversely, the DOMINO trial (n=1990 pregnant women) reported no difference in 1-hr post-OGTT glucose concentrations between DHA supplemented mothers and controls (74). The reason for such discrepancies is unclear but may be that n-3 PUFAs (such as DHA) require interactions with other metabolites (e.g., Vitamin D) (75) to impart an effect, concentrations of which vary considerably between populations, seasons, and geographic region (76-78).

The study aimed to increase and test generalizability of results within a diverse population; however, our results may not be generalizable across other ethnic groups or geographic regions. Nonetheless, this study has four main limitations. Firstly, samples were taken at a single time point before 28 weeks gestation, therefore (i) we were unable to account for differences in fasting duration and diurnal variation; and (ii) our results are not generalizable across the full-term of pregnancy. Secondly, as with all observational studies, the effect of confounding cannot be disregarded and causality cannot be inferred. Despite this, this is the first study to use a panel of multivariate statistical techniques to characterize GDM within a large prospective cohort with an equal representation of WE women and women from a non-WE population, meaning that statistical power to measure the same effect size is comparable between groups. Thirdly, the biological validity of the identified metabolites was tested and many correlated with postprandial glucose measures; and although confounding cannot be eliminated, all models included known GDM confounders and modelling characterizing the overall metabolic differences between ethnicities were also performed to test whether differences in metabolite profiles were found between ethnicities in relation to GDM status. Finally, diet is a contributor to metabolite concentrations, but comprehensive dietary data was not available for our analysis. Future work with comprehensive dietary records are needed evaluate the presence of a moderating effect of diet on metabolism and GDM risk.

## Conclusion

In conclusion, this study has identified unique and shared metabolic profiles that characterize GDM in WE and SA women. Future work exploring the moderating role of lifestyle on the metabolome and the underlying biological mechanisms driving the identified associations will provide a better understanding of the etiological factors responsible for the heightened level of GDM risk experienced by SA women and shed light on improved prevention strategies

## Data Availability

All data produced in the present work are contained in the manuscript.

## Abbreviations

ApoA1: Apolipoprotein A1
BIB: Born in Bradford
DHA: Docosahexanoic Acid
FAw3: Total Omega-3
FAw6: Total Omega-6
GDM: Gestational Diabetes Mellitus
GRM: Glycolysis Related Metabolites
HDL-C: High Density Lipoprotein Cholesterol
HD2L-C: High Density Lipoprotein-2 Cholesterol
IQR: Inter Quartile Range
LDL_D: Diameter of Low Density Lipoprotein
LPS: Lipoprotein Particle Size
MSEP: Mean Square Error of Prediction
MFA: Monounsaturated Fatty Acids
MW: Mann-Whitney
NHS: National Health Service
PCA: Principal Component Analyses
OGTT: Oral Glucose Tolerance Test
oPLSDA: Orthogonal Partial Least Squares Discriminatory Analyses
SAC-H: High Weight South Asian Cases
SANC-H: High Weight South Asian Non-Cases
SAC-N: Healthy Weight South Asian Cases
SANC-N: Healthy Weight South Asian Non-Cases
PLSDA: Partial Least Squares Discriminatory Analyses
RMSEE: Root Mean Square Error of Prediction
ROC: Receiver Operator Curve
SA: South Asian
SE: Standard Error
SFA: Saturated Fatty Acids
sPLSDA: Sparse Partial Least Squares Discriminatory Analyses
TCA: Tricarboxylic Acid Cycle
VIP: Variable Importance in Projection
VLDL_D: Diameter of Very Low Density Lipoprotein
WEs: White Europeans
WEC-N: Healthy Weight White European Cases
WENC-N: Healthy Weight White European Non-Cases
WEC-H: High Weight White European Cases
WENC-H: High Weight White European Non-Cases

## Acknowledgements

Born in Bradford (BiB) is only possible because of the enthusiasm and commitment of the children and parents in BiB. We are grateful to all the participants, health professionals, schools and researchers who have made Born in Bradford happen

