## Supplementary for "Unique Metabolic Profiles Associate with Gestational Diabetes and Ethnicity in Low and High-Risk Women Living in the UK"

**Online Supplementary Material**

Supplementary table 7: Coefficients of variation (CV) of included metabolites within both ethnicities.	29

### Supplementary table 1: Mean Variables of Importance in Projection (VIPs) for GDM adjusted covariates within ethnically stratified Partial Least Squares Discriminatory (PLSDA) models of pregnant women (mean gestational age 26.7 weeks).^[1]^

|  | **Prediction of GDM** | | |
| --- | --- | --- | --- |
| **Covariate** | **WE** | **SA** | **MW** **P- value** |
| Age (years) | 5.99 (0.27) | 5.84 (0.17) | 0.53 |
| BMI (continuous) | 4.33 (0.22) | 7.06 (0.21) | ≤ 0.001 |
| Parity | 2.91 (0.14) | 2.72 (0.04) | 0.41 |
| Multiple Pregnancy | 1.59 (0.13) | 1.38 (0.09) | 0.10 |
| Smoking Status | 2.12 (0.11) | 1.31 (0.10) | ≤ 0.001 |

### Supplementary table 2: Mean variables importance in projection (VIPs) of metabolite measures within stratified partial least squares discriminatory analysis (PLSDA) of pregnant women (mean gestational age 26.7 weeks). ^1^

|  |  |  | **Prediction of Case Status** | | | **Prediction of Ethnicity** | | |
| --- | --- | --- | --- | --- | --- | --- | --- | --- |
| **Metabolite Class** | **Metabolite subclass** | **Metabolite** | **WE** | **SA** | **MW P- value** | **Cases** | **Non - Cases** | **MW P- value** |
| Lipoproteins | Chylomicrons & extremely large VLDL | Total Lipids | 0.26 (0.03) | 0.28 (0.04) | 0.86 | 0.41 (0.05) | 0.30 (0.00) | 0.29 |
| Lipoproteins | Chylomicrons & extremely large VLDL | Phospholipids | 0.09 (0.01) | 0.08 (0.01) | 0.53 | 0.19 (0.03) | 0.20 (0.00) | 0.03 |
| Lipoproteins | Chylomicrons & extremely large VLDL | Total Cholesterol | 0.13 (0.02) | 0.13 (0.02) | 0.99 | 0.25 (0.04) | 0.24 (0.00) | 0.09 |
| Lipoproteins | Chylomicrons & extremely large VLDL | Cholesterol Esters | 0.13 (0.02) | 0.10 (0.01) | 0.90 | 0.29 (0.06) | 0.28 (0.00) | 0.04 |
| Lipoproteins | Chylomicrons & extremely large VLDL | Free Cholesterol | 0.07 (0.01) | 0.07 (0.01) | 0.76 | 0.18 (0.04) | 0.19 (0.00) | 0.03 |
| Lipoproteins | Chylomicrons & extremely large VLDL | Triglycerides | 0.20 (0.02) | 0.23 (0.04) | 0.97 | 0.39 (0.06) | 0.30 (0.00) | 0.11 |
| Lipoproteins | Very large VLDL | Total Lipids | 0.39 (0.03) | 0.30 (0.02) | 0.34 | 0.47 (0.05) | 0.37 (0.00) | 0.002 |
| Lipoproteins | Very large VLDL | Phospholipids | 0.18 (0.03) | 0.13 (0.01) | 0.51 | 0.30 (0.05) | 0.29 (0.00) | 0.02 |
| Lipoproteins | Very large VLDL | Total Cholesterol | 0.20 (0.02) | 0.20 (0.02) | 0.95 | 0.35 (0.05) | 0.27 (0.00) | 0.11 |
| Lipoproteins | Very large VLDL | Cholesterol Esters | 0.15 (0.01) | 0.16 (0.02) | 0.76 | 0.34 (0.06) | 0.27 (0.00) | 0.11 |
| Lipoproteins | Very large VLDL | Free Cholesterol | 0.16 (0.03) | 0.13 (0.02) | 0.64 | 0.25 (0.04) | 0.22 (0.00) | 0.03 |
| Lipoproteins | Very large VLDL | Triglycerides | 0.28 (0.02) | 0.23 (0.01) | 0.09 | 0.44 (0.06) | 0.35 (0.00) | 0.17 |
| Lipoproteins | Large VLDL | Total Lipids | 0.60 (0.01) | 0.49 (0.02) | 0.01 | 0.67 (0.04) | 0.59 (0.00) | ≤0.001 |
| Lipoproteins | Large VLDL | Phospholipids | 0.25 (0.01) | 0.22 (0.02) | 0.01 | 0.36 (0.04) | 0.34 (0.00) | 0.03 |
| Lipoproteins | Large VLDL | Total Cholesterol | 0.30 (0.02) | 0.27 (0.02) | 0.13 | 0.41 (0.04) | 0.34 (0.00) | 0.82 |
| Lipoproteins | Large VLDL | Cholesterol Esters | 0.22 (0.02) | 0.20 (0.01) | 0.50 | 0.37 (0.04) | 0.33 (0.00) | 0.11 |
| Lipoproteins | Large VLDL | Free Cholesterol | 0.23 (0.02) | 0.17 (0.01) | 0.03 | 0.35 (0.04) | 0.30 (0.00) | 0.01 |
| Lipoproteins | Large VLDL | Triglycerides | 0.49 (0.01) | 0.37 (0.01) | 0.01 | 0.56 (0.04) | 0.47 (0.00) | ≤0.001 |
| Lipoproteins | Medium VLDL | Total Lipids | 0.66 (0.05) | 0.56 (0.02) | ≤0.01 | 0.76 (0.04) | 0.69 (0.00) | ≤0.001 |
| Lipoproteins | Medium VLDL | Phospholipids | 0.28 (0.03) | 0.22 (0.01) | ≤0.01 | 0.40 (0.04) | 0.39 (0.00) | 0.03 |
| Lipoproteins | Medium VLDL | Total Cholesterol | 0.40 (0.04) | 0.38 (0.03) | 0.09 | 0.49 (0.03) | 0.45 (0.00) | 0.25 |
| Lipoproteins | Medium VLDL | Cholesterol Esters | 0.41 (0.05) | 0.42 (0.05) | 0.88 | 0.50 (0.05) | 0.40 (0.00) | 0.47 |
| Lipoproteins | Medium VLDL | Free Cholesterol | 0.24 (0.02) | 0.18 (0.01) | ≤0.01 | 0.36 (0.04) | 0.32 (0.00) | 0.11 |

|  |  |  | **Prediction of Case Status** | | | **Prediction of Ethnicity** | | |
| --- | --- | --- | --- | --- | --- | --- | --- | --- |
| **Metabolite Class** | **Metabolite subclass** | **Metabolite** | **WE** | **SA** | **MW P- value** | **Cases** | **Non - Cases** | **MW P- value** |
| Lipoproteins | Medium VLDL | Triglycerides | 0.54 (0.04) | 0.41 (0.01) | 0.01 | 0.60 (0.04) | 0.50 (0.00) | ≤0.001 |
| Lipoproteins | Small VLDL | Total Lipids | 0.62 (0.05) | 0.45 (0.03) | ≤0.01 | 0.69 (0.04) | 0.64 (0.00) | ≤0.001 |
| Lipoproteins | Small VLDL | Phospholipids | 0.43 (0.05) | 0.22 (0.01) | ≤0.01 | 0.41 (0.03) | 0.35 (0.00) | 0.01 |
| Lipoproteins | Small VLDL | Total Cholesterol | 0.45 (0.0) | 0.35 (0.03) | 0.11 | 0.49 (0.03) | 0.48 (0.00) | 0.66 |
| Lipoproteins | Small VLDL | Cholesterol Esters | 0.37 (0.04) | 0.31 (0.03) | 0.55 | 0.50 (0.04) | 0.43 (0.00) | 0.68 |
| Lipoproteins | Small VLDL | Free Cholesterol | 0.28 (0.03) | 0.17 (0.01) | ≤0.01 | 0.33 (0.03) | 0.30 (0.00) | 0.49 |
| Lipoproteins | Small VLDL | Triglycerides | 0.42 (0.04) | 0.35 (0.03) | 0.04 | 0.55 (0.05) | 0.44 (0.00) | 0.01 |
| Lipoproteins | Very small VLDL | Total Lipids | 0.59 (0.05) | 0.44 (0.03) | ≤0.01 | 0.65 (0.03) | 0.65 (0.00) | 0.29 |
| Lipoproteins | Very small VLDL | Phospholipids | 0.44 (0.06) | 0.26 (0.02) | 0.06 | 0.53 (0.05) | 0.42 (0.00) | 0.43 |
| Lipoproteins | Very small VLDL | Total Cholesterol | 0.52 (0.05) | 0.36 (0.03) | 0.02 | 0.52 (0.04) | 0.47 (0.00) | 0.56 |
| Lipoproteins | Very small VLDL | Cholesterol Esters | 0.56 (0.06) | 0.29 (0.02) | 0.07 | 0.55 (0.05) | 0.41 (0.00) | 0.09 |
| Lipoproteins | Very small VLDL | Free Cholesterol | 0.27 (0.04) | 0.19 (0.02) | 0.24 | 0.43 (0.05) | 0.37 (0.00) | 0.60 |
| Lipoproteins | Very small VLDL | Triglycerides | 0.37 (0.04) | 0.27 (0.02) | 0.18 | 0.49 (0.06) | 0.40 (0.00) | 0.23 |
| Lipoproteins | LDL | Concentration | 0.00 (0.00) | 0.00 (0.00) | 0.12 | 0.00 (0.00) | 0.04 (0.01) | ≤0.001 |
| Lipoproteins | LDL | Total Lipids | 0.62 (0.05) | 0.48 (0.02) | ≤0.01 | 0.81 (0.04) | 0.93 (0.00) | 0.01 |
| Lipoproteins | LDL | Phospholipids | 0.35 (0.03) | 0.25 (0.01) | ≤0.01 | 0.51 (0.04) | 0.49 (0.00) | 0.47 |
| Lipoproteins | LDL | Total Cholesterol | 0.56 (0.05) | 0.40 (0.02) | ≤0.01 | 0.69 (0.04) | 0.74 (0.00) | 0.06 |
| Lipoproteins | LDL | Cholesterol Esters | 0.55 (0.05) | 0.34 (0.01) | ≤0.01 | 0.62 (0.04) | 0.65 (0.00) | 0.04 |
| Lipoproteins | LDL | Free Cholesterol | 0.31 (0.04) | 0.21 (0.01) | ≤0.01 | 0.40 (0.03) | 0.42 (0.00) | 0.01 |
| Lipoproteins | LDL | Triglycerides | 0.40 (0.04) | 0.30 (0.01) | 0.12 | 0.44 (0.04) | 0.39 (0.00) | 0.82 |
| Lipoproteins | Large LDL | Concentration | 0.00 (0.00) | 0.00 (0.00) | ≤0.01 | 0.00 (0.00) | 0.03 (0.01) | ≤0.001 |
| Lipoproteins | Large LDL | Total Lipids | 0.70 (0.50) | 0.51 (0.02) | ≤0.01 | 0.85 (0.04) | 1.00 (0.00) | ≤0.001 |
| Lipoproteins | Large LDL | Phospholipids | 0.33 (0.03) | 0.23 (0.01) | ≤0.01 | 0.49 (0.04) | 0.49 (0.00) | 0.01 |
| Lipoproteins | Large LDL | Total Cholesterol | 0.62 (0.05) | 0.45 (0.02) | ≤0.01 | 0.73 (0.04) | 0.84 (0.00) | ≤0.001 |
| Lipoproteins | Large LDL | Cholesterol Esters | 0.55 (0.04) | 0.39 (0.02) | ≤0.01 | 0.65 (0.03) | 0.75 (0.00) | ≤0.001 |
| Lipoproteins | Large LDL | Free Cholesterol | 0.30 (0.03) | 0.23 (0.01) | ≤0.01 | 0.42 (0.03) | 0.43 (0.00) | 0.06 |
| Lipoproteins | Large LDL | Triglycerides | 0.38 (0.04) | 0.28 (0.02) | 0.07 | 0.44 (0.04) | 0.37 (0.00) | 0.16 |
| Lipoproteins | Medium LDL | Concentration | 0.00 (0.00) | 0.00 (0.00) | 0.01 | 0.00 (0.00) | 0.02 (0.00) | ≤0.001 |
| Lipoproteins | Medium LDL | Total Lipids | 0.58 (0.04) | 0.42 (0.02) | ≤0.01 | 0.68 (0.04) | 0.79 (0.00) | ≤0.001 |
| Lipoproteins | Medium LDL | Phospholipids | 0.26 (0.03) | 0.17 (0.01) | ≤0.01 | 0.40 (0.04) | 0.41 (0.00) | 0.01 |
| Lipoproteins | Medium LDL | Total Cholesterol | 0.51 (0.04) | 0.40 (0.02) | ≤0.01 | 0.59 (0.03) | 0.67 (0.00) | 0.004 |

|  |  |  | **Prediction of Case Status** | | | **Prediction of Ethnicity** | | |
| --- | --- | --- | --- | --- | --- | --- | --- | --- |
| **Metabolite Class** | **Metabolite subclass** | **Metabolite** | **WE** | **SA** | **MW P- value** | **Cases** | **Non - Cases** | **MW P- value** |
| Lipoproteins | Medium LDL | Cholesterol Esters | 0.48 (0.04) | 0.36 (0.02) | ≤0.01 | 0.56 (0.03) | 0.61 (0.00) | 0.04 |
| Lipoproteins | Medium LDL | Free Cholesterol | 0.25 (0.02) | 0.17 (0.01) | ≤0.01 | 0.34 (0.03) | 0.37 (0.00) | 0.01 |
| Lipoproteins | Medium LDL | Triglycerides | 0.31 (0.03) | 0.20 (0.01) | 0.09 | 0.35 (0.04) | 0.33 (0.00) | 0.09 |
| Lipoproteins | Small LDL | Concentration | 0.00 (0.00) | 0.00 (0.00) | ≤0.01 | 0.00 (0.00) | 0.03 (0.01) | ≤0.001 |
| Lipoproteins | Small LDL | Total Lipids | 0.47 (0.04) | 0.33 (0.01) | ≤0.01 | 0.58 (0.03) | 0.64 (0.00) | 0.02 |
| Lipoproteins | Small LDL | Phospholipids | 0.27 (0.03) | 0.17 (0.01) | ≤0.01 | 0.39 (0.04) | 0.37 (0.00) | 0.10 |
| Lipoproteins | Small LDL | Total Cholesterol | 0.42 (0.04) | 0.31 (0.01) | ≤0.01 | 0.49 (0.03) | 0.54 (0.00) | 0.03 |
| Lipoproteins | Small LDL | Cholesterol Esters | 0.38 (0.03) | 0.28 (0.01) | ≤0.01 | 0.44 (0.02) | 0.48 (0.00) | 0.05 |
| Lipoproteins | Small LDL | Free Cholesterol | 0.22 (0.02) | 0.16 (0.01) | 0.01 | 0.34 (0.04) | 0.29 (0.00) | 0.13 |
| Lipoproteins | Small LDL | Triglycerides | 0.19 (0.02) | 0.14 (0.01) | 0.02 | 0.31 (0.04) | 0.29 (0.00) | 0.05 |
| Lipoproteins | Extra-large HDL | Concentration | 0.00 (0.00) | 0.00 (0.00) | 0.10 | 0.00 (0.00) | 0.06 (0.01) | ≤0.001 |
| Lipoproteins | Extra-large HDL | Total Lipids | 0.81 (0.06) | 0.64 (0.03) | ≤0.01 | 0.80 (0.04) | 0.49 (0.00) | ≤0.001 |
| Lipoproteins | Extra-large HDL | Phospholipids | 0.67 (0.06) | 0.46 (0.03) | 0.05 | 0.66 (0.05) | 0.38 (0.00) | ≤0.001 |
| Lipoproteins | Extra-large HDL | Total Cholesterol | 0.63 (0.05) | 0.51 (0.02) | 0.06 | 0.64 (0.04) | 0.39 (0.00) | ≤0.001 |
| Lipoproteins | Extra-large HDL | Cholesterol Esters | 0.59 (0.05) | 0.41 (0.02) | 0.17 | 0.56 (0.04) | 0.37 (0.00) | ≤0.001 |
| Lipoproteins | Extra-large HDL | Free Cholesterol | 0.35 (0.03) | 0.26 (0.01) | 0.16 | 0.41 (0.04) | 0.27 (0.00) | ≤0.001 |
| Lipoproteins | Extra-large HDL | Triglycerides | 0.23 (0.02) | 0.17 (0.01) | 0.80 | 0.34 (0.05) | 0.31 (0.00) | 0.03 |
| Lipoproteins | Large HDL | Concentration | 0.00 (0.00) | 0.00 (0.00) | 0.02 | 0.00 (0.00) | 0.08 (0.01) | ≤0.001 |
| Lipoproteins | Large HDL | Total Lipids | 0.86 (0.06) | 0.67 (0.03) | ≤0.01 | 0.86 (0.05) | 0.44 (0.01) | ≤0.001 |
| Lipoproteins | Large HDL | Phospholipids | 0.59 (0.05) | 0.42 (0.02) | 0.02 | 0.67 (0.06) | 0.39 (0.00) | ≤0.001 |
| Lipoproteins | Large HDL | Total Cholesterol | 0.67 (0.05) | 0.51 (0.02) | 0.01 | 0.65 (0.04) | 0.35 (0.00) | ≤0.001 |
| Lipoproteins | Large HDL | Total Cholesterol | 0.67 (0.05) | 0.51 (0.02) | 0.01 | 0.65 (0.04) | 0.35 (0.00) | ≤0.001 |
| Lipoproteins | Large HDL | Cholesterol Esters | 0.60 (0.05) | 0.44 (0.02) | 0.01 | 0.57 (0.04) | 0.33 (0.00) | ≤0.001 |
| Lipoproteins | Large HDL | Free Cholesterol | 0.33 (0.03) | 0.23 (0.01) | 0.05 | 0.35 (0.03) | 0.21 (0.00) | ≤0.001 |
| Lipoproteins | Large HDL | Triglycerides | 0.40 (0.06) | 0.23 (0.03) | 0.58 | 0.41 (0.06) | 0.32 (0.00) | 0.29 |
| Lipoproteins | Medium HDL | Concentration | 0.00 (0.00) | 0.00 (0.00) | 0.01 | 0.00 (0.00) | 0.09 (0.01) | ≤0.001 |
| Lipoproteins | Medium HDL | Total Lipids | 0.91 (0.08) | 0.74 (0.03) | ≤0.01 | 0.79 (0.04) | 0.28 (0.00) | ≤0.001 |
| Lipoproteins | Medium HDL | Phospholipids | 0.66 (0.06) | 0.50 (0.02) | 0.01 | 0.64 (0.06) | 0.30 (0.00) | ≤0.001 |
| Lipoproteins | Medium HDL | Total Cholesterol | 0.72 (0.05) | 0.56 (0.02) | ≤0.01 | 0.63 (0.04) | 0.28 (0.00) | ≤0.001 |
| Lipoproteins | Medium HDL | Cholesterol Esters | 0.68 (0.05) | 0.49 (0.02) | 0.01 | 0.60 (0.04) | 0.28 (0.00) | ≤0.001 |
| Lipoproteins | Medium HDL | Free Cholesterol | 0.33 (0.03) | 0.24 (0.01) | 0.01 | 0.32 (0.03) | 0.20 (0.00) | ≤0.001 |

|  |  |  | **Prediction of Case Status** | | | **Prediction of Ethnicity** | | |
| --- | --- | --- | --- | --- | --- | --- | --- | --- |
| **Metabolite Class** | **Metabolite subclass** | **Metabolite** | **WE** | **SA** | **MW P- value** | **Cases** | **Non - Cases** | **MW P- value** |
| Lipoproteins | Medium HDL | Triglycerides | 0.37 (0.04) | 0.27 (0.03) | 0.66 | 0.42 (0.05) | 0.31 (0.00) | 0.21 |
| Lipoproteins | Small HDL | Concentration | 0.00 (0.00) | 0.00 (0.00) | 0.01 | 0.00 (0.00) | 0.09 (0.01) | ≤0.001 |
| Lipoproteins | Small HDL | Total Lipids | 0.97 (0.07) | 0.77 (0.02) | ≤0.01 | 0.79 (0.05) | 0.30 (0.00) | ≤0.001 |
| Lipoproteins | Small HDL | Phospholipids | 0.79 (0.06) | 0.62 (0.03) | ≤0.01 | 0.63 (0.05) | 0.32 (0.00) | ≤0.001 |
| Lipoproteins | Small HDL | Total Cholesterol | 0.71 (0.05) | 0.55 (0.02) | 0.01 | 0.61 (0.04) | 0.31 (0.00) | ≤0.001 |
| Lipoproteins | Small HDL | Cholesterol Esters | 0.69 (0.05) | 0.51 (0.01) | 0.05 | 0.60 (0.04) | 0.32 (0.00) | ≤0.001 |
| Lipoproteins | Small HDL | Free Cholesterol | 0.37 (0.04) | 0.24 (0.01) | 0.02 | 0.41 (0.06) | 0.26 (0.00) | 0.13 |
| Lipoproteins | Small HDL | Triglycerides | 0.28 (0.03) | 0.19 (0.01) | 0.13 | 0.35 (0.05) | 0.34 (0.00) | 0.01 |
| Lipoproteins | Lipoprotein particle size | Mean diameter VLDL | 1.58 (0.07) | 1.38 (0.07) | 0.04 | 1.51 (0.09) | 0.88 (0.00) | ≤0.001 |
| Lipoproteins | Lipoprotein particle size | Mean diameter LDL | 0.94 (0.07) | 0.88 (0.04) | 0.58 | 0.92 (0.06) | 0.36 (0.00) | ≤0.001 |
| Lipoproteins | Lipoprotein particle size | Mean diameter HDL | 0.87 (0.06) | 0.75 (0.04) | 0.03 | 0.85 (0.05) | 0.44 (0.00) | ≤0.001 |
| Amino Acids | Unbranched | Alanine | 1.02 (0.1) | 0.89 (0.05) | 0.09 | 1.10 (0.08) | 0.35 (0.00) | ≤0.001 |
| Amino Acids | Unbranched | Glutamine | 1.08 (0.09) | 0.96 (0.06) | 0.18 | 0.74 (0.08) | 0.34 (0.01) | ≤0.001 |
| Amino Acids | Unbranched | Glycine | 0.89 (0.10) | 0.73 (0.07) | 0.231 | 0.70 (0.06) | 0.39 (0.00) | ≤0.001 |
| Amino Acids | Unbranched | Histidine | 0.55 (0.08) | 0.79 (0.05) | 0.02 | 0.49 (0.06) | 0.36 (0.00) | 0.10 |
| Amino Acids | Branched chain | Isoleucine | 0.53 (0.08) | 0.51 (0.06) | 0.99 | 0.46 (0.06) | 0.32 (0.00) | 0.13 |
| Amino Acids | Branched chain | Leucine | 0.59 (0.09) | 0.60 (0.07) | 0.97 | 0.51 (0.06) | 0.33 (0.00) | 0.01 |
| Amino Acids | Branched chain | Valine | 0.71 (0.08) | 0.77 (0.06) | 0.66 | 0.56 (0.06) | 0.31 (0.00) | 0.003 |
| Amino Acids | Aromatic | Phenylalanine | 0.89 (0.07) | 0.53 (0.07) | ≤0.01 | 0.72 (0.06) | 0.39 (0.00) | ≤0.001 |
| Amino Acids | Aromatic | Tyrosine | 0.61 (0.09) | 0.51 (0.06) | 0.37 | 0.43 (0.05) | 0.39 0.01) | 0.29 |
| Apolipoproteins | Apolipoproteins | Apolipoprotein A1 | 0.70 (0.01) | 0.54 (0.01) | ≤0.01 | 0.73 (0.05) | 1.12 (0.00) | ≤0.001 |
| Apolipoproteins | Apolipoproteins | Apolipoprotein B | 0.57 (0.05) | 0.37 (0.02) | ≤0.01 | 0.81 (0.06) | 0.54 (0.00) | 0.01 |
| Cholesterol | Cholesterol | Total serum cholesterol | 1.04 (0.04) | 0.71 (0.03) | ≤0.01 | 1.33 (0.07) | 1.69 (0.01) | ≤0.001 |
| Cholesterol | Cholesterol | VLDL cholesterol | 0.62 (0.02) | 0.48 (0.03) | ≤0.01 | 0.74 (0.04) | 0.80 (0.00) | 0.05 |
| Cholesterol | Cholesterol | Remnant cholesterol | 0.75 (0.02) | 0.54 (0.03) | ≤0.01 | 0.92 (0.05) | 1.10 (0.00) | ≤0.001 |
| Cholesterol | Cholesterol | LDL Cholesterol | 0.84 (0.02) | 0.64 (0.02) | ≤0.01 | 1.01 (0.05) | 1.17 (0.00) | 0.01 |
| Cholesterol | Cholesterol | HDL Cholesterol | 0.80 (0.04) | 0.62 (0.02) | ≤0.01 | 0.81 (0.04) | 0.46 (0.01) | ≤0.001 |
| Cholesterol | Cholesterol | HDL2 Cholesterol | 0.80 (0.03) | 0.62 (0.01) | ≤0.01 | 0.78 (0.04) | 0.43 (0.01) | ≤0.001 |
| Cholesterol | Cholesterol | HDL3 Cholesterol | 0.56 (0.07) | 0.39 (0.04) | 0.36 | 0.56 (0.06) | 0.39 (0.00) | 0.03 |
| Cholesterol | Cholesterol | Total esterified cholesterol | 0.99 (0.03) | 0.76 (0.04) | ≤0.01 | 1.18 (0.06) | 1.42 (0.01) | ≤0.001 |

|  |  |  | **Prediction of Case Status** | | | **Prediction of Ethnicity** | | |
| --- | --- | --- | --- | --- | --- | --- | --- | --- |
| **Metabolite Class** | **Metabolite subclass** | **Metabolite** | **WE** | **SA** | **MW P- value** | **Cases** | **Non - Cases** | **MW P- value** |
| Cholesterol | Cholesterol | Total free Cholesterol | 0.78 (0.05) | 0.66 (0.04) | 0.06 | 0.87 (0.05) | 0.98 (0.00) | 0.01 |
| Fatty Acids | Fatty Acids | Total fatty acids | 1.71 (0.09) | 1.13 (0.05) | ≤0.01 | 2.15 (0.12) | 0.83 (0.01) | ≤0.001 |
| Fatty Acids | Fatty Acids | Docosahexaenoic acid | 0.75 (0.07) | 0.65 (0.05) | 0.18 | 0.79 (0.05) | 2.71 (0.00) | ≤0.001 |
| Fatty Acids | Fatty Acids | 18:2 Linoleic acid | 1.34 (0.06) | 1.24 (0.03) | 0.11 | 1.38 (0.07) | 0.37 (0.01) | ≤0.001 |
| Fatty Acids | Fatty Acids | n-3 fatty acids | 0.85 (0.08) | 0.73 (0.06) | 0.17 | 1.03 (0.06) | 1.26 (0.00) | ≤0.001 |
| Fatty Acids | Fatty Acids | n-6 fatty acids | 1.08 (0.03) | 0.92 (0.03) | ≤0.01 | 1.28 (0.07) | 0.49 (0.01) | 0.13 |
| Fatty Acids | Fatty Acids | PUFA | 1.08 (0.03) | 0.92 (0.03) | ≤0.01 | 1.33 (0.07) | 1.42 (0.01) | 0.12 |
| Fatty Acids | Fatty Acids | MUFA | 1.60 (0.04) | 1.22 (0.05) | ≤0.01 | 1.49 (0.08) | 1.48 (0.00) | 0.02 |
| Fatty Acids | Fatty Acids | SFA | 1.40 (0.06) | 1.00 (0.05) | ≤0.01 | 1.51 (0.08) | 1.52 (0.01) | 0.06 |
| Glycerides and Phospholipids | Glycerides and Phospholipids | Total serum triglycerides | 0.88 (0.02) | 0.67 (0.01) | ≤0.01 | 0.98 (0.05) | 0.99 (0.00) | 0.11 |
| Glycerides and Phospholipids | Glycerides and Phospholipids | VLDL triglycerides | 0.85 (0.01) | 0.65 (0.01) | ≤0.01 | 0.89 (0.05) | 0.81 (0.00) | ≤0.001 |
| Glycerides and Phospholipids | Glycerides and Phospholipids | LDL triglycerides | 0.49 (0.03) | 0.39 (0.03) | 0.03 | 0.52 (0.03) | 0.47 (0.00) | 0.03 |
| Glycerides and Phospholipids | Glycerides and Phospholipids | HDL triglycerides | 0.44 (0.04) | 0.30 (0.02) | 0.13 | 0.49 (0.05) | 0.41 (0.00) | 0.82 |
| Glycerides and Phospholipids | Glycerides and Phospholipids | Phosphoglycerides | 0.96 (0.04) | 0.72 (0.03) | ≤0.01 | 1.02 (0.06) | 1.01 (0.00) | 0.03 |
| Glycerides and Phospholipids | Glycerides and Phospholipids | Phosphatidylcholine | 0.92 (0.04) | 0.70 (0.04) | ≤0.01 | 0.98 (0.05) | 1.12 (0.00) | 0.002 |
| Glycerides and Phospholipids | Glycerides and Phospholipids | Sphingomyelins | 0.69 (0.06) | 0.56 (0.06) | 0.18 | 0.65 (0.05) | 0.54 (0.00) | 0.02 |
| Glycerides and Phospholipids | Glycerides and Phospholipids | Total cholines | 0.96 (0.04) | 0.74 (0.04) | ≤0.01 | 1.08 (0.06) | 1.12 (0.00) | 0.68 |
| Glycolysis Related Metabolites | Glycolysis Related Metabolites | Lactate | 1.86 (0.17) | 1.73 (0.05) | 0.19 | 1.60 (0.09) | 0.53 (0.00) | ≤0.001 |
| Glycolysis Related Metabolites | Glycolysis Related Metabolites | Pyruvate | 0.79 (0.09) | 0.62 (0.05) | 0.06 | 0.66 (0.06) | 0.28 (0.00) | ≤0.001 |
| Glycolysis Related Metabolites | Glycolysis Related Metabolites | Citrate | 1.12 (0.09) | 0.57 (0.06) | ≤0.01 | 0.91 (0.07) | 0.29 (0.00) | ≤0.001 |

|  |  |  | **Prediction of Case Status** | | | **Prediction of Ethnicity** | | |
| --- | --- | --- | --- | --- | --- | --- | --- | --- |
| **Metabolite Class** | **Metabolite subclass** | **Metabolite** | **WE** | **SA** | **MW** **P- value** | **Cases** | **Non - Cases** | **MW P- value** |
| Glycolysis Related Metabolites | Glycolysis Related Metabolites | Glycerol | 0.69 (0.07) | 0.60 (0.05) | 0.43 | 0.53 (0.08) | 0.34 (0.00) | ≤0.001 |
| Ketone Bodies | Ketone Bodies | Acetate | 0.59 (0.09) | 0.59 (0.07) | 0.84 | 0.50 (0.05) | 0.28 (0.00) | ≤0.001 |
| Ketone Bodies | Ketone Bodies | Beta- hydroxybutyrate | 0.85 (0.10) | 0.87 (0.04) | 0.29 | 0.72 (0.07) | 0.35 (0.00) | ≤0.001 |
| Fluid Balance and Inflammation | Fluid Balance and Inflammation | Creatinine | 0.64 (0.09) | 0.51 (0.06) | 0.51 | 0.66 (0.05) | 0.27 (0.00) | ≤0.001 |
| Fluid Balance and Inflammation | Fluid Balance and Inflammation | Albumin | 0.65 (0.07) | 0.91 (0.05) | 0.01 | 0.85 (0.06) | 0.35 (0.00) | ≤0.001 |
| Fluid Balance and Inflammation | Fluid Balance and Inflammation | Glycoprotein acetyls | 1.25 (0.08) | 1.09 (0.06) | 0.13 | 1.14 (0.07) | 0.53 (0.01) | ≤0.001 |

^1^ VIPS were mean averaged across 20 iterations of PLSDA models and presented alongside their standard errors (SE) shown in brackets. All model iterations were significant (p value R^2^<0.05, p value Q^2^ < 0.05). The distributions of VIP values for each metabolite measure were compared between populations using a MW test. Sample sizes for each analysis were as followed: prediction of GDM status within WEs: 256; prediction of GDM status within SAs: 572; prediction of ethnicity within cases: 256; prediction of ethnicity within non-cases: 4770. GDM, Gestational Diabetes Mellitus; MW, Mann-Whitney; SA, South Asian; WE, white European.

|  | **Optimised component number** | | **All components** | |
| --- | --- | --- | --- | --- |
| **Iteration** | **White European** | **South Asian** | **White European** | **South Asian** |
| 1 | 0.572 | 0.247 | 0.621 | 0.364 |
| 2 | 0.363 | 0.149 | 0.651 | 0.298 |
| 3 | 0.631 | 0.325 | 0.656 | 0.336 |
| 4 | 0.301 | 0.210 | 0.594 | 0.353 |
| 5 | 0.201 | 0.177 | 0.564 | 0.364 |
| 6 | 0.477 | 0.194 | 0.602 | 0.352 |
| 7 | 0.370 | 0.155 | 0.609 | 0.353 |
| 8 | 0.168 | 0.157 | 0.555 | 0.348 |
| 9 | 0.316 | 0.186 | 0.591 | 0.334 |
| 10 | 0.285 | 0.131 | 0.547 | 0.301 |
| 11 | 0.187 | 0.182 | 0.604 | 0.332 |
| 12 | 0.350 | 0.236 | 0.636 | 0.365 |
| 13 | 0.106 | 0.336 | 0.609 | 0.354 |
| 14 | 0.234 | 0.243 | 0.586 | 0.378 |
| 15 | 0.194 | **0.338** | 0.553 | 0.368 |
| 16 | 0.234 | 0.203 | 0.583 | 0.343 |
| 17 | 0.227 | 0.188 | 0.601 | 0.302 |
| 18 | 0.146 | 0.392 | 0.631 | 0.403 |
| 19 | 0.218 | 0.243 | 0.619 | 0.297 |
| 20 | 0.421 | 0.242 | 0.596 | 0.347 |
| **Median (range)** | **0.26 (0.11 – 0.63)** | **0.20 (0.13- 0.39)** | **0.60 (0.55-0.66)** | 0.35 **(0.30–0.40)** |

### Supplementary table 3: Table of cumulative R^2^Y values for Partial Least Squares Discriminatory Analyses (PLSDA) predicting GDM case status in distinct ethnic strata from the Born in Bradford (BiB) cohort. ^[2]^

### Supplementary table 4: VIPs for the prediction of ethnicity within the overall Born in Bradford population via Partial Least Squares Discriminatory Analysis (PLSDA). ^[3]^

| **Variable** | **Model 1** | **Model 2** |
| --- | --- | --- |
| Age | 5.49 | 5.34 |
| Smoking Status | 5.08 | 4.83 |
| Parity | 5.01 | 4.83 |
| BMI | 4.68 | 4.56 |
| Total Fatty Acids | 2.60 | 2.55 |
| Serum Cholesterol | 1.60 | 1.58 |
| SFA | 1.56 | 1.55 |
| MUFA | 1.42 | 1.42 |
| PUFA | 1.41 | 1.40 |
| FAw6 | 1.35 | 1.34 |
| GDM Status | - | 1.33 |
| Esterified Cholesterol | 1.33 | 1.33 |
| 18:2 Linoleic Acid | 1.18 | 1.19 |
| LDL Cholesterol | 1.10 | 1.09 |
| Remnant Cholesterol | 1.04 | 1.03 |
| Phosphatidylchlorine | 1.03 | 1.05 |
| Total Cholesterol | 1.03 | 1.04 |

### Supplementary figure 1: Circular bar plot of metabolite variables of importance in projection (VIPs) from Partial Least Squares Discriminatory Analyses (PLSDA) models predicting ethnicity in GDM cases and non-GDM women.

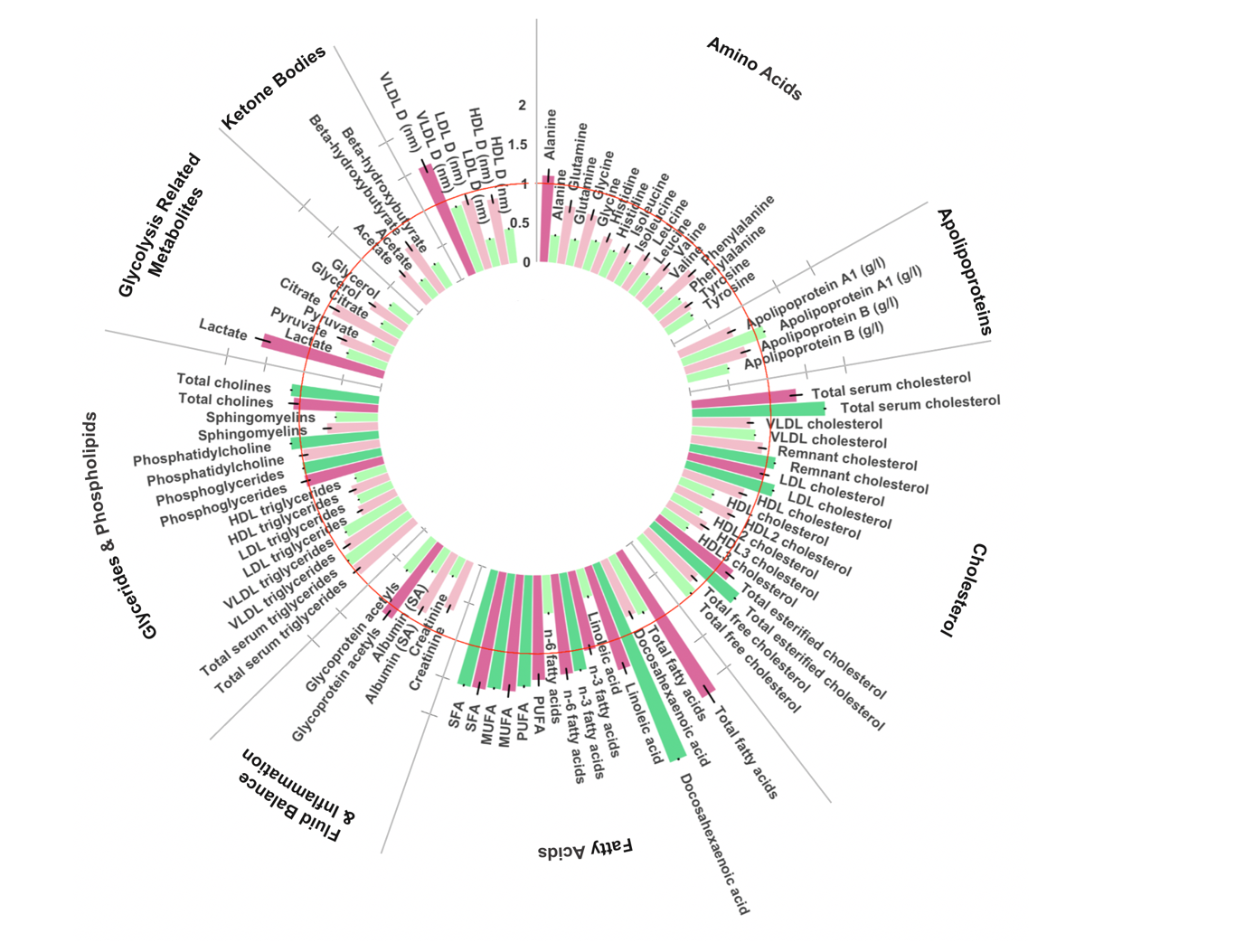

Circular bar plot of VIPS from 20 iterations of PLSDA models predicting ethnicity in cases (n=256) and non-cases (n= 4770). Bars represent standard errors. PLSDA adjusted for maternal age (years), BMI (continuous), smoking status, parity, and multiple pregnancy status. Red line denotes VIP cut-off of 1. Dark pink: GDM cases, VIP ≥ 1, light pink: GDM cases, VIP <1, dark green: GDM non-cases, VIP ≥1, light green: GDM non-case, VIP <1. Units mmol/L unless stated. GRM: Glycolysis Related Metabolites; LPS: Lipoprotein Particle Size; MUFA: total monounsaturated fatty acids; SFA: total saturated fatty acids; VLDL_D: mean diameter of very-low density lipoprotei

### Supplementary figure 2: GDM in Low-Risk Women (non-smokers with no previous children below the age of 35) characterised by sparse Partial Least Squares Discriminatory Analyses (sPLSDA).

**A**

**B**

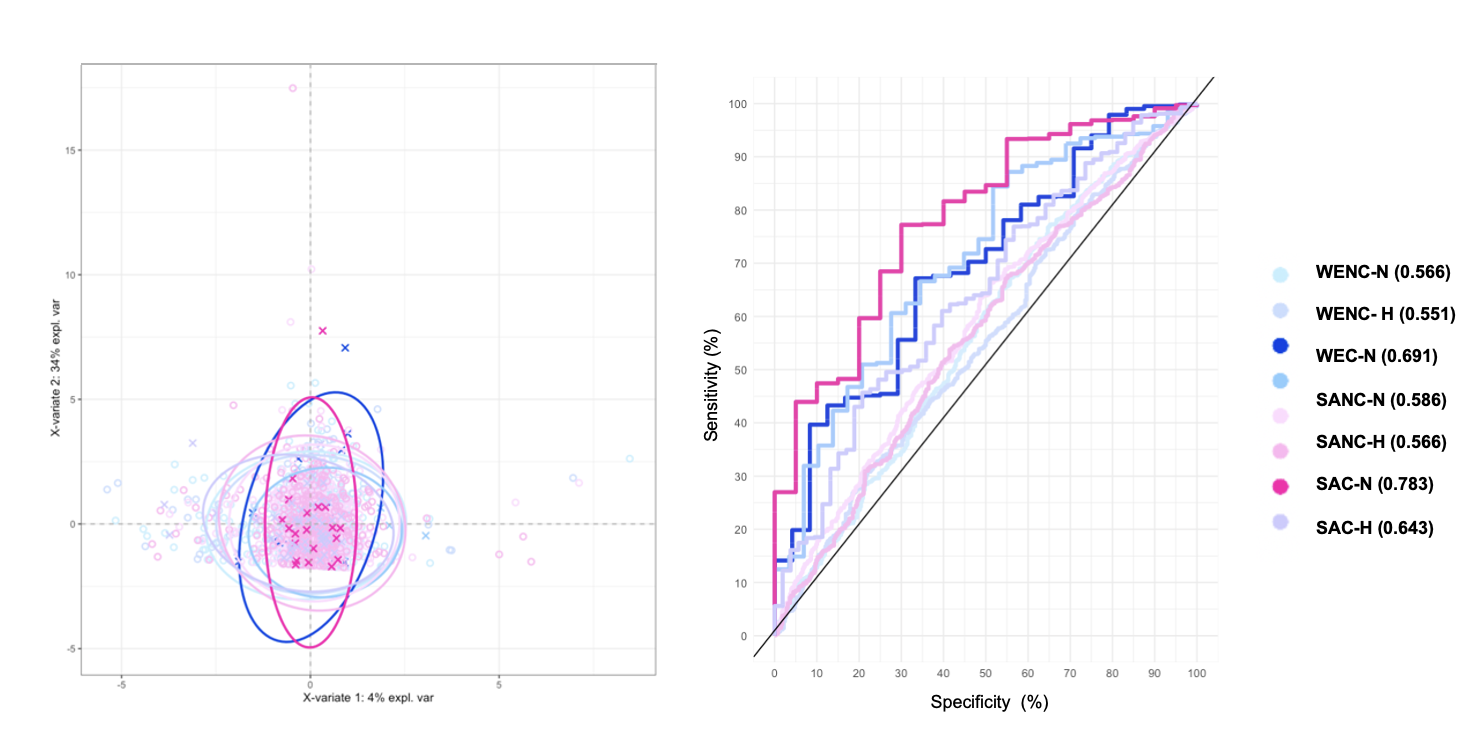

**A:** sPLSDA plot for the separation of low-risk mothers (n=1385) based upon their ethnicity, GDM status and BMI (normal vs high). **B**. Receiver Operator Curve (ROC) for sPLSDA model shown in **A**. SAC-H: high weight South Asian case (n=53) ; SAC-N: healthy weight South Asian case (n=20) ; SANC-H: high weight South Asian non-case (n=384); SANC-N health weight South Asian non case (n=407); WENC-H: high weight White European non-case (n=29) ; WEC-N: healthy weight White European Case (n=24); WENC-H: high weight White European non-case (n=374); WENC-N: healthy weight White European non-case (n=445).

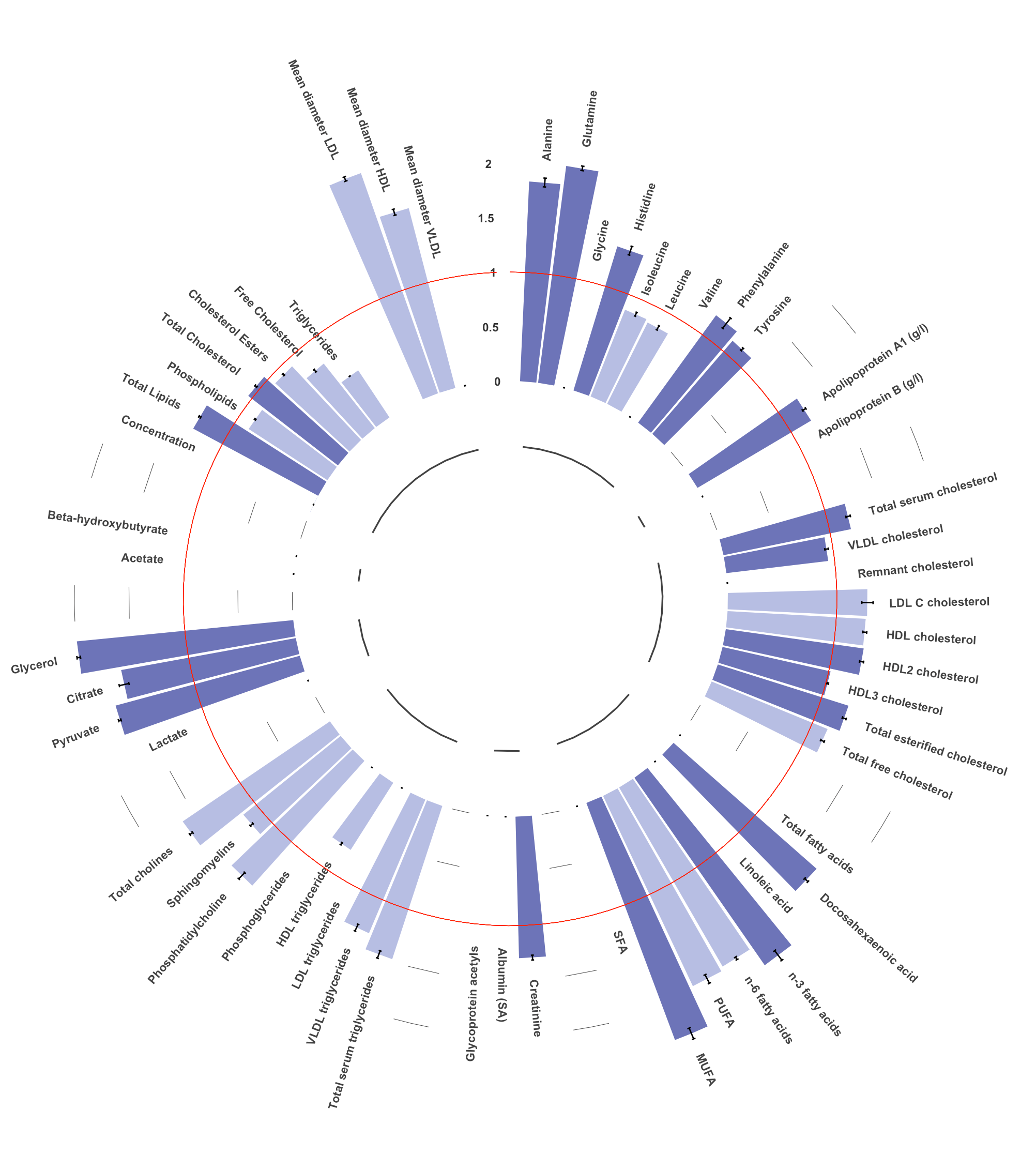

### Supplementary figure 3: Circular bar plots for variables of importance in projection (VIPs) from Partial Least Squares Discriminatory Analyses (PLSDA) of metabolites identified in sparse Partial Least Squares Discriminatory Analyses (sPLSDA) to be important in distinguishing healthy-weight South Asian cases.

**A**

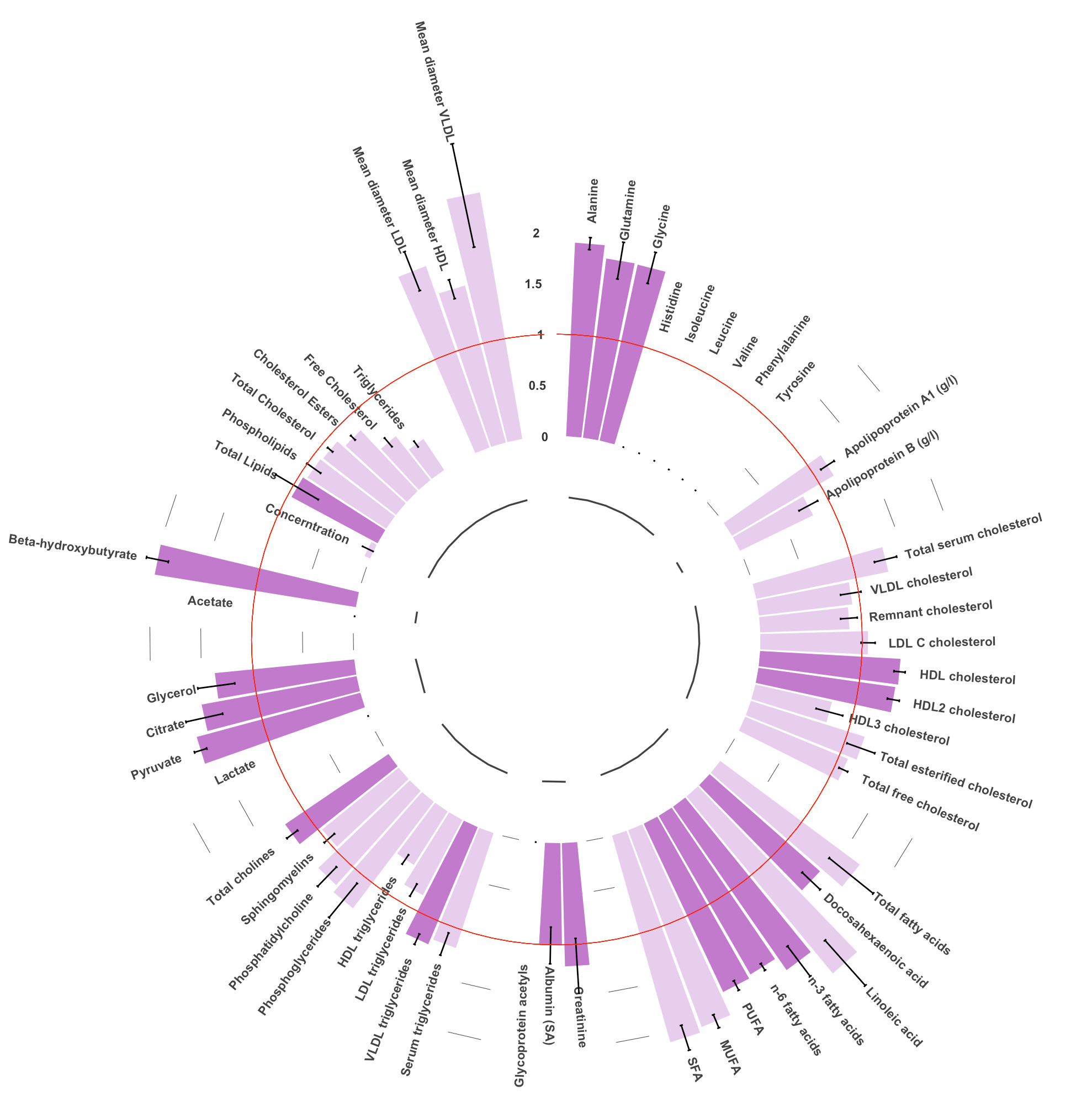

**B**

Mean

**C**

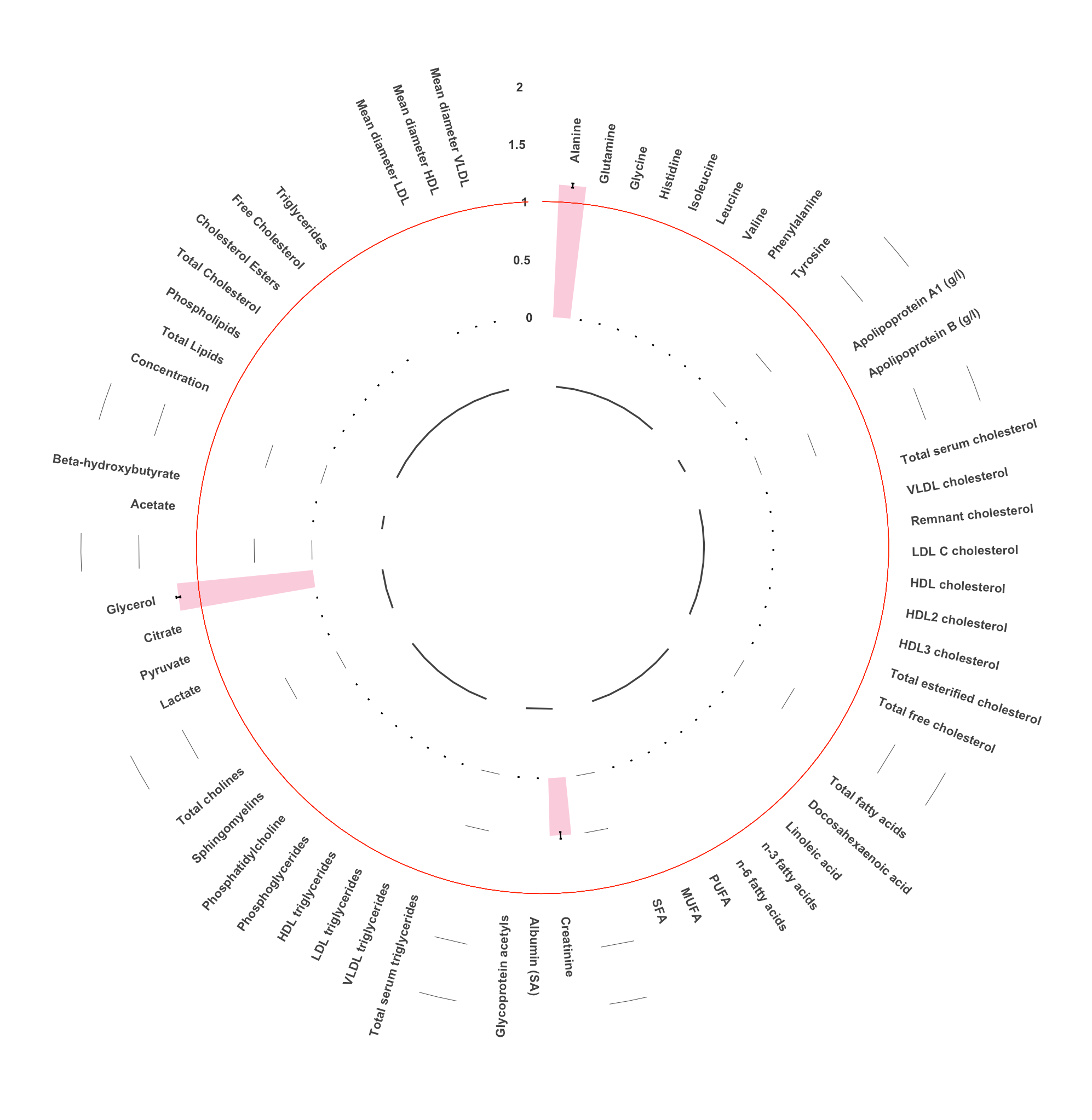

Mean VIPs and standard errors of metabolites driving the distinction between **A**: healthy-weight SA cases vs healthy-weight WE cases. **B**: healthy-weight SA cases vs high-weight SA cases **C**: healthy-weight SA cases from healthy-weight SA non-cases. Light Shaded colours illustrate metabolites selected based upon their high correlation (Pearson’s correlation ≥ 0.90) with a metabolite identified within sPLSDA. Metabolite measures absent from the circle plot/ with a VIP of 0 indicate metabolite measures which were not identified as important in sPLSDA. All units mmol/L unless stated. SA: Surface area.

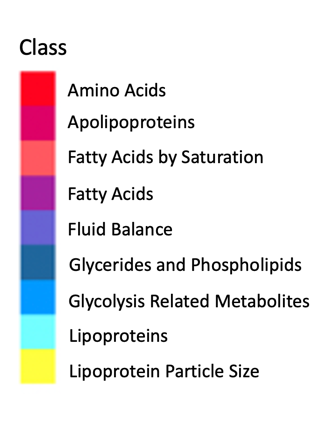

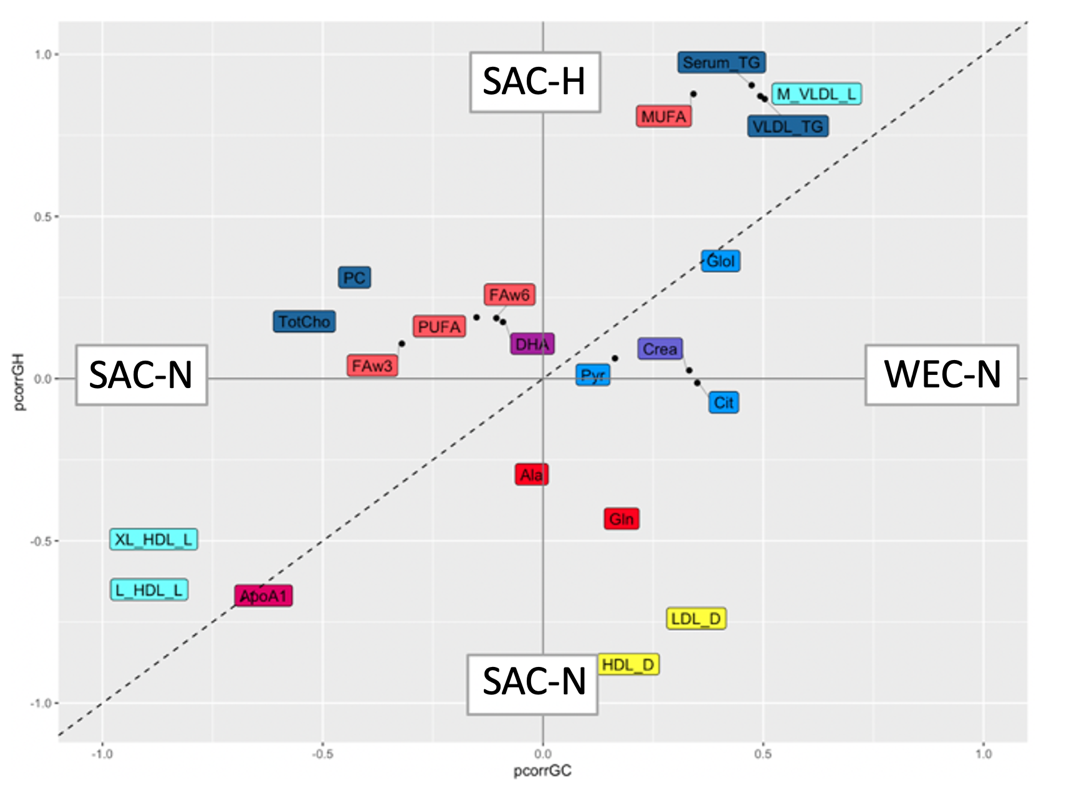

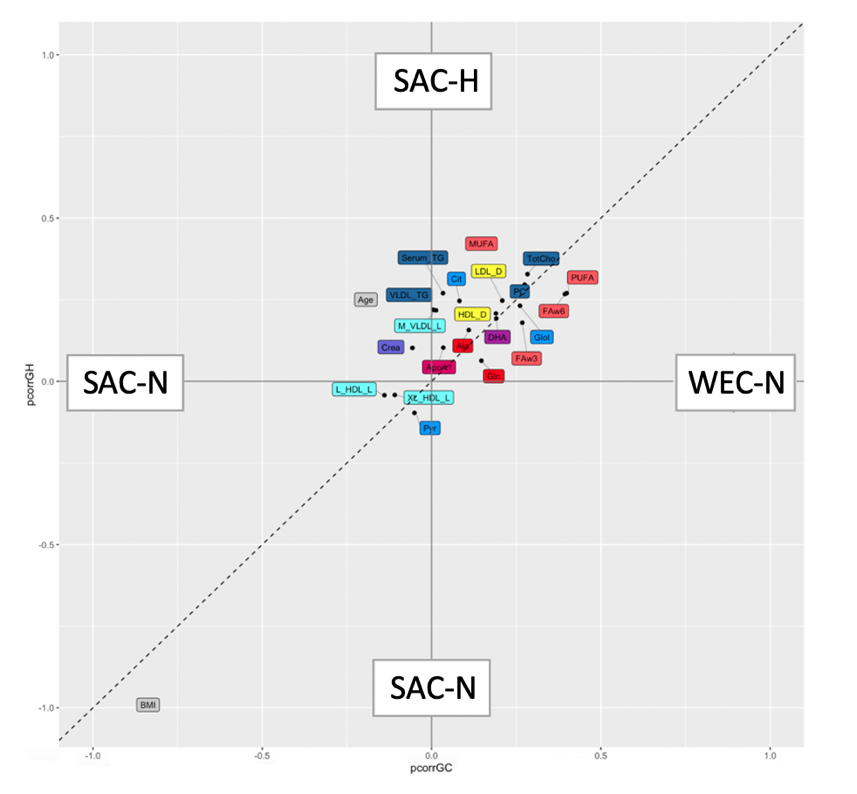

### Supplementary figure 4: Shared and Unique Structure (SUS) plots of shared metabolite determinants identified in sparse Partial Least Squares Discriminatory Analyses (sPLDSA) models characterising healthy weight South Asian cases (SAC-N), healthy weight South Asian non-cases (SANC-N), high weight South Asian non-cases and healthy weight white European non-cases (WEC-N)

Distance along the diagonals represents higher reliability. Distance along the horizontal represents higher magnitude. Top right/ bottom left corner represents metabolite values with the higher magnitude and higher reliability. A: Model including 34 metabolites identified within sPLSDA analysis. Orthogonal PLSDA (oPLSDA) models were non-significant. **B**: Model A + adjustment for maternal age and BMI (continuous). Center metabolites represent lowest magnitude and lower reliability. OPLS-DA p-value SAC-N vs SAC-H p-value< 0.05, SAC-N vs WEC-N p-value < 0.05.

**A**

**B**

### Supplementary table 5: Linear Regression of identified metabolites on log fasting glucose concentration (mmol/l) within pregnant women before 28 weeks’ gestation of distinct ethnicities from the Born in Bradford cohort. ^1^

**A**

**A**

|  | **Overall (n=5538*)** | | | | | | | | |
| --- | --- | --- | --- | --- | --- | --- | --- | --- | --- |
|  | **Model 1** | | | **Model 2** | | | **Model 3** | | |
| **Metabolite Measure** | **β** | **SE** | **P value** | **β** | **SE** | **P value** | **β** | **SE** | **P value** |
| Lactate (mmol/l) | -0.008 | 0.003 | 0.003 | -0.009 | 0.003 | 0.001 | -0.009 | 0.003 | 0.001 |
| Mean Diameter for VLDL particles (nm) | 0.000 | 0.001 | 0.824 |  |  |  |  |  |  |
| Total Fatty Acids (mmol/l) | 0.000 | 0.000 | 0.718 |  |  |  |  |  |  |
| MUFA (mmol/l) | 0.001 | 0.001 | 0.507 |  |  |  |  |  |  |
| 18:2 Linoleic Acid (mmol/l) | 0.000 | 0.002 | 0.892 |  |  |  |  |  |  |
| SFA (mmol/l) | 0.000 | 0.001 | 0.770 |  |  |  |  |  |  |
| Esterified Cholesterol (mmol/l) | 0.001 | 0.002 | 0.706 |  |  |  |  |  |  |
| Analine (mmol/l) | -0.034 | 0.029 | 0.247 |  |  |  |  |  |  |
| Glutamine (mmol/l) | -0.036 | 0.031 | 0.243 |  |  |  |  |  |  |
| Total Serum Cholesterol | 0.000 | 0.001 | 0.725 |  |  |  |  |  |  |
| n-6 Fatty Acids (mmol/l) | 0.000 | 0.002 | 0.999 |  |  |  |  |  |  |
| PUFA (mmol/l) | 0.000 | 0.001 | 0.965 |  |  |  |  |  |  |
| Glycoprotein Acetyls (mmol/l) | 0.005 | 0.007 | 0.488 |  |  |  |  |  |  |
| Citrate (mmol/l) | -0.071 | 0.082 | 0.388 |  |  |  |  |  |  |
| Glycine (mmol/l) | -0.055 | 0.043 | 0.203 |  |  |  |  |  |  |
| Histidine (mmol/l) | -0.055 | 0.047 | 0.239 |  |  |  |  |  |  |
| Phenylanaline (mmol/l) | -0.016 | 0.032 | 0.624 |  |  |  |  |  |  |
| Tyrosine (mmol/l) | -0.030 | 0.041 | 0.461 |  |  |  |  |  |  |
| Apolipoprotein A1 (g/l) | -0.009 | 0.006 | 0.166 |  |  |  |  |  |  |
| LDL Cholesterol (mmol/l) | 0.002 | 0.002 | 0.514 |  |  |  |  |  |  |
| HDL Cholesterol (mmol/l) | -0.006 | 0.004 | 0.142 |  |  |  |  |  |  |
| HDL2 Cholesterol (mmol/l) | -0.006 | 0.004 | 0.134 |  |  |  |  |  |  |
| HDL3 Cholesterol (mmol/l) | -0.030 | 0.032 | 0.343 |  |  |  |  |  |  |
| n-3 Fatty Acids (mmol/l) | 0.005 | 0.013 | 0.694 |  |  |  |  |  |  |
| DHA (mmol/l) | 0.031 | 0.031 | 0.230 |  |  |  |  |  |  |
| Creatine (mmol/l) | -0.368 | 0.226 | 0.103 |  |  |  |  |  |  |
| Albumin (Signal Area) | -0.599 | 0.238 | 0.012 | -0.596 | 0.231 | 0.010 | -0.611 | 0.233 | 0.009 |
| Serum triglycerides (mmol/l) | 0.002 | 0.002 | 0.445 |  |  |  |  |  |  |
| VLDL triglycerides (mmol/l) | 0.002 | 0.003 | 0.537 |  |  |  |  |  |  |
| Phosphoglycerides (mmol/l) | -0.001 | 0.003 | 0.751 |  |  |  |  |  |  |
| Phosphatidylchlorine (mmol/l) | 0.001 | 0.003 | 0.861 |  |  |  |  |  |  |
| Sphingomyelins (mmol/l) | -0.002 | 0.016 | 0.897 |  |  |  |  |  |  |
| Total Cholines (mmol/l) | 0.000 | 0.003 | 0.991 |  |  |  |  |  |  |
| Pyruvate (mmol/l) | -0.008 | 0.064 | 0.899 |  |  |  |  |  |  |
| Glycerol (mmol/l) | 0.000 | 0.086 | 0.996 |  |  |  |  |  |  |
| Beta- hydroxybutyrate (mmol/l) | 0.068 | 0.035 | 0.057 |  |  |  |  |  |  |
| Mean Diameter for LDL particles (nm) | 0.051 | 0.022 | 0.020 | 0.047 | 0.021 | 0.027 | 0.049 | 0.021 | 0.022 |
| Mean Diameter for HDL particles (nm) | 0.005 | 0.007 | 0.474 |  |  |  |  |  |  |

| **B**  **A** | **White European (n=2267)*** | | | | | | | | |
| --- | --- | --- | --- | --- | --- | --- | --- | --- | --- |
|  | **Model 1** | | | **Model 2** | | | **Model 3** | | |
| **Metabolite Measure** | **β** | **SE** | **P value** | **β** | **SE** | **P value** | **β** | **SE** | **P value** |
| Lactate (mmol/l) | -0.009 | 0.003 | 0.006 | -0.011 | 0.003 | 0.001 | -0.010 | 0.003 | 0.003 |
| Mean Diameter for VLDL particles (nm) | 0.001 | 0.002 | 0.545 |  |  |  |  |  |  |
| Total Fatty Acids (mmol/l) | 0.000 | 0.001 | 0.926 |  |  |  |  |  |  |
| MUFA (mmol/l) | 0.000 | 0.002 | 0.917 |  |  |  |  |  |  |
| 18:2 Linoleic Acid (mmol/l) | -0.001 | 0.002 | 0.784 |  |  |  |  |  |  |
| SFA (mmol/l) | 0.000 | 0.001 | 0.995 |  |  |  |  |  |  |
| Esterified Cholesterol (mmol/l) | -0.001 | 0.002 | 0.575 |  |  |  |  |  |  |
| Analine (mmol/l) | -0.014 | 0.034 | 0.674 |  |  |  |  |  |  |
| Glutamine (mmol/l) | -0.036 | 0.036 | 0.315 |  |  |  |  |  |  |
| Total Serum Cholesterol | -0.001 | 0.001 | 0.580 |  |  |  |  |  |  |
| n-6 Fatty Acids (mmol/l) | -0.001 | 0.002 | 0.652 |  |  |  |  |  |  |
| PUFA (mmol/l) | -0.001 | 0.002 | 0.698 |  |  |  |  |  |  |
| Glycoprotein Acetyls (mmol/l) | 0.011 | 0.009 | 0.211 |  |  |  |  |  |  |
| Citrate (mmol/l) | -0.087 | 0.095 | 0.362 |  |  |  |  |  |  |
| Glycine (mmol/l) | 0.060 | 0.050 | 0.225 |  |  |  |  |  |  |
| Histidine (mmol/l) | -0.428 | 0.173 | 0.014 | -0.459 | 0.167 | 0.006 | -0.449 | 0.170 | 0.008 |
| Phenylanaline (mmol/l) | -0.065 | 0.151 | 0.666 |  |  |  |  |  |  |
| Tyrosine (mmol/l) | -0.373 | 0.303 | 0.219 |  |  |  |  |  |  |
| Apolipoprotein A1 (g/l) | -0.018 | 0.007 | 0.014 | -0.017 | 0.007 | 0.019 | -0.018 | 0.007 | 0.014 |
| LDL Cholesterol (mmol/l) | 0.000 | 0.003 | 0.955 |  |  |  |  |  |  |
| HDL Cholesterol (mmol/l) | -0.011 | 0.004 | 0.012 | -0.010 | 0.004 | 0.019 | -0.011 | 0.004 | 0.011 |
| HDL2 Cholesterol (mmol/l) | -0.013 | 0.005 | 0.011 | -0.011 | 0.005 | 0.019 | -0.012 | 0.005 | 0.010 |
| HDL3 Cholesterol (mmol/l) | -0.066 | 0.037 | 0.077 |  |  |  |  |  |  |
| n-3 Fatty Acids (mmol/l) | 0.003 | 0.015 | 0.830 |  |  |  |  |  |  |
| DHA (mmol/l) | 0.042 | 0.036 | 0.249 |  |  |  |  |  |  |
| Creatine (mmol/l) | -0.166 | 0.277 | 0.548 |  |  |  |  |  |  |
| Albumin (Signal Area) | -0.387 | 0.276 | 0.160 |  |  |  |  |  |  |
| Serum triglycerides (mmol/l) | 0.002 | 0.003 | 0.433 |  |  |  |  |  |  |
| VLDL triglycerides (mmol/l) | 0.003 | 0.004 | 0.416 |  |  |  |  |  |  |
| Phosphoglycerides (mmol/l) | -0.004 | 0.004 | 0.313 |  |  |  |  |  |  |
| Phosphatidylchlorine (mmol/l) | -0.002 | 0.003 | 0.548 |  |  |  |  |  |  |
| Sphingomyelins (mmol/l) | -0.016 | 0.019 | 0.381 |  |  |  |  |  |  |
| Total Cholines (mmol/l) | -0.002 | 0.003 | 0.458 |  |  |  |  |  |  |
| Pyruvate (mmol/l) | 0.113 | 0.075 | 0.134 |  |  |  |  |  |  |
| Glycerol (mmol/l) | 0.075 | 0.101 | 0.457 |  |  |  |  |  |  |
| Beta- hydroxybutyrate (mmol/l) | 0.070 | 0.039 | 0.074 |  |  |  |  |  |  |
| Mean Diameter for LDL particles (nm) | 0.061 | 0.025 | 0.016 | 0.055 | 0.024 | 0.025 | 0.056 | 0.025 | 0.024 |
| Mean Diameter for HDL particles (nm) | -0.002 | 0.008 | 0.783 |  |  |  |  |  |  |

| **C**  **A** | **South Asian (n=2671)** | | | | | | | | |
| --- | --- | --- | --- | --- | --- | --- | --- | --- | --- |
|  | **Model 1** | | | **Model 2** | | | **Model 3** | | |
| **Metabolite Measure** | **β** | **SE** | **P value** | **β** | **SE** | **P value** | **β** | **SE** | **P value** |
| Lactate (mmol/l) | -0.007 | 0.004 | 0.105 |  |  |  |  |  |  |
| Mean Diameter for VLDL particles (nm) | -0.001 | 0.002 | 0.561 |  |  |  |  |  |  |
| Total Fatty Acids (mmol/l) | 0.000 | 0.001 | 0.605 |  |  |  |  |  |  |
| MUFA (mmol/l) | 0.002 | 0.002 | 0.435 |  |  |  |  |  |  |
| 18:2 Linoleic Acid (mmol/l) | 0.001 | 0.003 | 0.683 |  |  |  |  |  |  |
| SFA (mmol/l) | 0.001 | 0.002 | 0.726 |  |  |  |  |  |  |
| Esterified Cholesterol (mmol/l) | 0.002 | 0.003 | 0.510 |  |  |  |  |  |  |
| Analine (mmol/l) | -0.042 | 0.046 | 0.360 |  |  |  |  |  |  |
| Glutamine (mmol/l) | -0.024 | 0.049 | 0.619 |  |  |  |  |  |  |
| Total Serum Cholesterol | 0.001 | 0.002 | 0.526 |  |  |  |  |  |  |
| n-6 Fatty Acids (mmol/l) | 0.001 | 0.003 | 0.752 |  |  |  |  |  |  |
| PUFA (mmol/l) | 0.001 | 0.002 | 0.745 |  |  |  |  |  |  |
| Glycoprotein Acetyls (mmol/l) | 0.003 | 0.011 | 0.772 |  |  |  |  |  |  |
| Citrate (mmol/l) | -0.031 | 0.131 | 0.813 |  |  |  |  |  |  |
| Glycine (mmol/l) | -0.029 | 0.069 | 0.676 |  |  |  |  |  |  |
| Histidine (mmol/l) | -0.044 | 0.054 | 0.417 |  |  |  |  |  |  |
| Phenylanaline (mmol/l) | -0.017 | 0.036 | 0.633 |  |  |  |  |  |  |
| Tyrosine (mmol/l) | -0.031 | 0.046 | 0.498 |  |  |  |  |  |  |
| Apolipoprotein A1 (g/l) | -0.002 | 0.010 | 0.875 |  |  |  |  |  |  |
| LDL Cholesterol (mmol/l) | 0.002 | 0.004 | 0.501 |  |  |  |  |  |  |
| HDL Cholesterol (mmol/l) | -0.001 | 0.006 | 0.824 |  |  |  |  |  |  |
| HDL2 Cholesterol (mmol/l) | -0.002 | 0.007 | 0.808 |  |  |  |  |  |  |
| HDL3 Cholesterol (mmol/l) | -0.001 | 0.051 | 0.983 |  |  |  |  |  |  |
| n-3 Fatty Acids (mmol/l) | 0.007 | 0.020 | 0.745 |  |  |  |  |  |  |
| DHA (mmol/l) | 0.035 | 0.049 | 0.474 |  |  |  |  |  |  |
| Creatine (mmol/l) | -0.619 | 0.343 | 0.071 |  |  |  |  |  |  |
| Albumin (Signal Area) | -0.866 | 0.382 | 0.024 | -0.818 | 0.366 | 0.025 | -0.858 | 0.375 | 0.022 |
| Serum triglycerides (mmol/l) | 0.002 | 0.004 | 0.660 |  |  |  |  |  |  |
| VLDL triglycerides (mmol/l) | 0.001 | 0.005 | 0.792 |  |  |  |  |  |  |
| Phosphoglycerides (mmol/l) | 0.002 | 0.005 | 0.766 |  |  |  |  |  |  |
| Phosphatidylchlorine (mmol/l) | 0.003 | 0.005 | 0.576 |  |  |  |  |  |  |
| Sphingomyelins (mmol/l) | 0.007 | 0.025 | 0.790 |  |  |  |  |  |  |
| Total Cholines (mmol/l) | 0.002 | 0.004 | 0.645 |  |  |  |  |  |  |
| Pyruvate (mmol/l) | -0.115 | 0.101 | 0.254 |  |  |  |  |  |  |
| Glycerol (mmol/l) | -0.080 | 0.137 | 0.559 |  |  |  |  |  |  |
| Beta- hydroxybutyrate (mmol/l) | 0.073 | 0.060 | 0.225 |  |  |  |  |  |  |
| Mean Diameter for LDL particles (nm) | 0.049 | 0.035 | 0.158 |  |  |  |  |  |  |
| Mean Diameter for HDL particles (nm) | 0.009 | 0.011 | 0.428 |  |  |  |  |  |  |

^1^ β coefficients of metabolite measures from linear regression models predicting fasting glucose status. * 1 white European sample had a missing fasting glucose measure **and** was excluded from the analysis. Model 1 adjusted for maternal age (years), gestational age (days), parity and smoking status during pregnancy (yes/no). When β coefficients were significant (P value ≤ 0.05) models were additionally adjusted for BMI as a continuous variable (Model 2) and BMI as a dichotomous variable, grouping mothers based on whether they were above their ethnic specific cut off for overweight on the BMI scale (i.e, ≥ 25 kg/m^2^ in white Europeans ≥ 23.5 kg/m^2^ in South Asians). (Model 3) **A**: Overall population results. **B**: white European. **C:** South Asian.

### Supplementary table 6: Linear Regression of identified metabolites on 2-hour post glucose concentration (mmol/l) within pregnant women before 28 weeks’ gestation of distinct ethnicities from the Born in Bradford cohort. ^1^

**A**

**A**

|  | **Overall (n=5538*)** | | | | | | | | |
| --- | --- | --- | --- | --- | --- | --- | --- | --- | --- |
|  | **Model 1** | | | **Model 2** | | | **Model 3** | | |
| **Metabolite Measure** | **β** | **SE** | **P value** | **β** | **SE** | **P value** | **β** | **SE** | **P value** |
| Lactate (mmol/l) | -0.008 | 0.007 | 0.246 |  |  |  |  |  |  |
| Mean Diameter for VLDL particles (nm) | 0.000 | 0.003 | 0.996 |  |  |  |  |  |  |
| Total Fatty Acids (mmol/l) | 0.001 | 0.001 | 0.285 |  |  |  |  |  |  |
| MUFA (mmol/l) | 0.004 | 0.003 | 0.239 |  |  |  |  |  |  |
| 18:2 Linoleic Acid (mmol/l) | 0.001 | 0.004 | 0.808 |  |  |  |  |  |  |
| SFA (mmol/l) | 0.004 | 0.003 | 0.221 |  |  |  |  |  |  |
| Esterified Cholesterol (mmol/l) | 0.005 | 0.004 | 0.243 |  |  |  |  |  |  |
| Analine (mmol/l) | 0.009 | 0.068 | 0.899 |  |  |  |  |  |  |
| Glutamine (mmol/l) | -0.052 | 0.072 | 0.471 |  |  |  |  |  |  |
| Total Serum Cholesterol | 0.003 | 0.003 | 0.267 |  |  |  |  |  |  |
| n-6 Fatty Acids (mmol/l) | 0.002 | 0.004 | 0.624 |  |  |  |  |  |  |
| PUFA (mmol/l) | 0.002 | 0.003 | 0.566 |  |  |  |  |  |  |
| Glycoprotein Acetyls (mmol/l) | 0.006 | 0.017 | 0.710 |  |  |  |  |  |  |
| Citrate (mmol/l) | 0.160 | 0.192 | 0.405 |  |  |  |  |  |  |
| Glycine (mmol/l) | -0.164 | 0.101 | 0.103 |  |  |  |  |  |  |
| Histidine (mmol/l) | -0.094 | 0.110 | 0.391 |  |  |  |  |  |  |
| Phenylanaline (mmol/l) | -0.048 | 0.074 | 0.521 |  |  |  |  |  |  |
| Tyrosine (mmol/l) | -0.074 | 0.095 | 0.437 |  |  |  |  |  |  |
| Apolipoprotein A1 (g/l) | 0.018 | 0.015 | 0.238 |  |  |  |  |  |  |
| LDL Cholesterol (mmol/l) | 0.004 | 0.005 | 0.479 |  |  |  |  |  |  |
| HDL Cholesterol (mmol/l) | 0.010 | 0.009 | 0.253 |  |  |  |  |  |  |
| HDL2 Cholesterol (mmol/l) | 0.010 | 0.010 | 0.289 |  |  |  |  |  |  |
| HDL3 Cholesterol (mmol/l) | 0.114 | 0.075 | 0.129 |  |  |  |  |  |  |
| n-3 Fatty Acids (mmol/l) | 0.033 | 0.030 | 0.276 |  |  |  |  |  |  |
| DHA (mmol/l) | 0.129 | 0.072 | 0.075 |  |  |  |  |  |  |
| Creatine (mmol/l) | 0.108 | 0.528 | 0.837 |  |  |  |  |  |  |
| Albumin (Signal Area) | -0.773 | 0.165 | 0.165 |  |  |  |  |  |  |
| Serum triglycerides (mmol/l) | 0.003 | 0.006 | 0.628 |  |  |  |  |  |  |
| VLDL triglycerides (mmol/l) | 0.001 | 0.007 | 0.931 |  |  |  |  |  |  |
| Phosphoglycerides (mmol/l) | 0.011 | 0.008 | 0.171 |  |  |  |  |  |  |
| Phosphatidylchlorine (mmol/l) | 0.009 | 0.007 | 0.199 |  |  |  |  |  |  |
| Sphingomyelins (mmol/l) | 0.031 | 0.037 | 0.408 |  |  |  |  |  |  |
| Total Cholines (mmol/l) | 0.009 | 0.007 | 0.164 |  |  |  |  |  |  |
| Pyruvate (mmol/l) | 0.110 | 0.150 | 0.463 |  |  |  |  |  |  |
| Glycerol (mmol/l) | 0.112 | 0.201 | 0.578 |  |  |  |  |  |  |
| Beta- hydroxybutyrate (mmol/l) | 0.006 | 0.083 | 0.945 |  |  |  |  |  |  |
| Mean Diameter for LDL particles (nm) | 0.058 | 0.051 | 0.253 |  |  |  |  |  |  |
| Mean Diameter for HDL particles (nm) | 0.022 | 0.016 | 0.158 |  |  |  |  |  |  |

**B**

**A**

|  | **White European (n=2267)*** | | | | | | | | |
| --- | --- | --- | --- | --- | --- | --- | --- | --- | --- |
|  | **Model 1** | | | **Model 2** | | | **Model 3** | | |
| **Metabolite Measure** | **β** | **SE** | **P value** | **β** | **SE** | **P value** | **β** | **SE** | **P value** |
| Lactate (mmol/l) | -0.008 | 0.009 | 0.380 |  |  |  |  |  |  |
| Mean Diameter for VLDL particles (nm) | 0.001 | 0.004 | 0.842 |  |  |  |  |  |  |
| Total Fatty Acids (mmol/l) | 0.002 | 0.002 | 0.181 |  |  |  |  |  |  |
| MUFA (mmol/l) | 0.005 | 0.004 | 0.249 |  |  |  |  |  |  |
| 18:2 Linoleic Acid (mmol/l) | 0.005 | 0.006 | 0.434 |  |  |  |  |  |  |
| SFA (mmol/l) | 0.006 | 0.004 | 0.118 |  |  |  |  |  |  |
| Esterified Cholesterol (mmol/l) | 0.010 | 0.006 | 0.075 |  |  |  |  |  |  |
| Analine (mmol/l) | 0.080 | 0.093 | 0.390 |  |  |  |  |  |  |
| Glutamine (mmol/l) | 0.021 | 0.098 | 0.833 |  |  |  |  |  |  |
| Total Serum Cholesterol | 0.007 | 0.004 | 0.091 |  |  |  |  |  |  |
| n-6 Fatty Acids (mmol/l) | 0.005 | 0.005 | 0.351 |  |  |  |  |  |  |
| PUFA (mmol/l) | 0.005 | 0.005 | 0.298 |  |  |  |  |  |  |
| Glycoprotein Acetyls (mmol/l) | 0.009 | 0.023 | 0.692 |  |  |  |  |  |  |
| Citrate (mmol/l) | -0.164 | 0.260 | 0.527 |  |  |  |  |  |  |
| Glycine (mmol/l) | -0.110 | 0.135 | 0.417 |  |  |  |  |  |  |
| Histidine (mmol/l) | -0.213 | 0.472 | 0.652 |  |  |  |  |  |  |
| Phenylanaline (mmol/l) | 0.011 | 0.411 | 0.979 |  |  |  |  |  |  |
| Tyrosine (mmol/l) | -0.598 | 0.824 | 0.468 |  |  |  |  |  |  |
| Apolipoprotein A1 (g/l) | 0.023 | 0.020 | 0.268 |  |  |  |  |  |  |
| LDL Cholesterol (mmol/l) | 0.011 | 0.007 | 0.126 |  |  |  |  |  |  |
| HDL Cholesterol (mmol/l) | 0.009 | 0.012 | 0.474 |  |  |  |  |  |  |
| HDL2 Cholesterol (mmol/l) | 0.008 | 0.013 | 0.534 |  |  |  |  |  |  |
| HDL3 Cholesterol (mmol/l) | 0.128 | 0.101 | 0.205 |  |  |  |  |  |  |
| n-3 Fatty Acids (mmol/l) | 0.069 | 0.041 | 0.097 |  |  |  |  |  |  |
| DHA (mmol/l) | 0.196 | 0.098 | 0.047 | 0.203 | 0.097 | 0.037 | 0.195 | 0.097 | 0.045 |
| Creatine (mmol/l) | -0.302 | 0.753 | 0.688 |  |  |  |  |  |  |
| Albumin (Signal Area) | 0.294 | 0.750 | 0.695 |  |  |  |  |  |  |
| Serum triglycerides (mmol/l) | 0.006 | 0.008 | 0.459 |  |  |  |  |  |  |
| VLDL triglycerides (mmol/l) | 0.005 | 0.010 | 0.630 |  |  |  |  |  |  |
| Phosphoglycerides (mmol/l) | 0.014 | 0.011 | 0.190 |  |  |  |  |  |  |
| Phosphatidylchlorine (mmol/l) | 0.013 | 0.009 | 0.167 |  |  |  |  |  |  |
| Sphingomyelins (mmol/l) | 0.074 | 0.051 | 0.151 |  |  |  |  |  |  |
| Total Cholines (mmol/l) | 0.013 | 0.009 | 0.147 |  |  |  |  |  |  |
| Pyruvate (mmol/l) | 0.193 | 0.205 | 0.348 |  |  |  |  |  |  |
| Glycerol (mmol/l) | 0.172 | 0.273 | 0.530 |  |  |  |  |  |  |
| Beta- hydroxybutyrate (mmol/l) | -0.077 | 0.106 | 0.468 |  |  |  |  |  |  |
| Mean Diameter for LDL particles (nm) | -0.077 | 0.106 | 0.468 |  |  |  |  |  |  |
| Mean Diameter for HDL particles (nm) | 0.021 | 0.021 | 0.328 |  |  |  |  |  |  |

**C**

**A**

|  | **South Asian (n=2671)** | | | | | | | | |
| --- | --- | --- | --- | --- | --- | --- | --- | --- | --- |
|  | **Model 1** | | | **Model 2** | | | **Model 3** | | |
| **Metabolite Measure** | **β** | **SE** | **P value** | **β** | **SE** | **P value** | **β** | **SE** | **P value** |
| Lactate (mmol/l) | -0.007 | 0.009 | 0.479 |  |  |  |  |  |  |
| Mean Diameter for VLDL particles (nm) | 0.000 | 0.005 | 0.979 |  |  |  |  |  |  |
| Total Fatty Acids (mmol/l) | 0.000 | 0.002 | 0.793 |  |  |  |  |  |  |
| MUFA (mmol/l) | 0.003 | 0.005 | 0.547 |  |  |  |  |  |  |
| 18:2 Linoleic Acid (mmol/l) | -0.002 | 0.006 | 0.718 |  |  |  |  |  |  |
| SFA (mmol/l) | 0.001 | 0.004 | 0.790 |  |  |  |  |  |  |
| Esterified Cholesterol (mmol/l) | -0.001 | 0.006 | 0.862 |  |  |  |  |  |  |
| Analine (mmol/l) | -0.043 | 0.098 | 0.665 |  |  |  |  |  |  |
| Glutamine (mmol/l) | -0.111 | 0.104 | 0.285 |  |  |  |  |  |  |
| Total Serum Cholesterol | -0.001 | 0.004 | 0.875 |  |  |  |  |  |  |
| n-6 Fatty Acids (mmol/l) | -0.001 | 0.006 | 0.856 |  |  |  |  |  |  |
| PUFA (mmol/l) | -0.001 | 0.005 | 0.869 |  |  |  |  |  |  |
| Glycoprotein Acetyls (mmol/l) | 0.010 | 0.024 | 0.690 |  |  |  |  |  |  |
| Citrate (mmol/l) | 0.511 | 0.281 | 0.069 |  |  |  |  |  |  |
| Glycine (mmol/l) | -0.196 | 0.148 | 0.187 |  |  |  |  |  |  |
| Histidine (mmol/l) | -0.101 | 0.116 | 0.384 |  |  |  |  |  |  |
| Phenylanaline (mmol/l) | -0.055 | 0.077 | 0.481 |  |  |  |  |  |  |
| Tyrosine (mmol/l) | -0.076 | 0.098 | 0.438 |  |  |  |  |  |  |
| Apolipoprotein A1 (g/l) | 0.010 | 0.022 | 0.652 |  |  |  |  |  |  |
| LDL Cholesterol (mmol/l) | -0.004 | 0.008 | 0.586 |  |  |  |  |  |  |
| HDL Cholesterol (mmol/l) | 0.010 | 0.013 | 0.458 |  |  |  |  |  |  |
| HDL2 Cholesterol (mmol/l) | 0.010 | 0.014 | 0.479 |  |  |  |  |  |  |
| HDL3 Cholesterol (mmol/l) | 0.090 | 0.109 | 0.408 |  |  |  |  |  |  |
| n-3 Fatty Acids (mmol/l) | 0.000 | 0.044 | 0.992 |  |  |  |  |  |  |
| DHA (mmol/l) | 0.071 | 0.105 | 0.497 |  |  |  |  |  |  |
| Creatine (mmol/l) | 0.365 | 0.734 | 0.619 |  |  |  |  |  |  |
| Albumin (Signal Area) | -1.900 | 0.816 | 0.020 | -1.824 | 0.797 | 0.022 | -1.887 | 0.808 | 0.020 |
| Serum triglycerides (mmol/l) | 0.001 | 0.008 | 0.940 |  |  |  |  |  |  |
| VLDL triglycerides (mmol/l) | -0.002 | 0.010 | 0.837 |  |  |  |  |  |  |
| Phosphoglycerides (mmol/l) | 0.008 | 0.012 | 0.504 |  |  |  |  |  |  |
| Phosphatidylchlorine (mmol/l) | 0.004 | 0.010 | 0.661 |  |  |  |  |  |  |
| Sphingomyelins (mmol/l) | -0.016 | 0.054 | 0.767 |  |  |  |  |  |  |
| Total Cholines (mmol/l) | 0.005 | 0.010 | 0.583 |  |  |  |  |  |  |
| Pyruvate (mmol/l) | 0.046 | 0.216 | 0.832 |  |  |  |  |  |  |
| Glycerol (mmol/l) | 0.042 | 0.292 | 0.885 |  |  |  |  |  |  |
| Beta- hydroxybutyrate (mmol/l) | 0.118 | 0.128 | 0.355 |  |  |  |  |  |  |
| Mean Diameter for LDL particles (nm) | 0.097 | 0.075 | 0.194 |  |  |  |  |  |  |
| Mean Diameter for HDL particles (nm) | 0.019 | 0.023 | 0.416 |  |  |  |  |  |  |

^1^ β coefficients of metabolite measures from linear regression models predicting fasting glucose status. Model 1 adjusted for maternal age (years), gestational age (days), parity and smoking status during pregnancy (yes/no). When β coefficients were significant (P value ≤ 0.05) models were additionally adjusted for BMI as a continuous variable (Model 2) and BMI as a dichotomous variable, grouping mothers based on whether they were above their ethnic specific cut off for overweight on the BMI scale. (Model 3) **A**: Overall sample results. **B**: White European. **C:** South Asian.

| **Metabolite**  Supplementary table 7: Coefficients of variation (CV) of included metabolites within both ethnicities. | **White European** | **South Asian** |
| --- | --- | --- |
| XXL_VLDL_P | 75.3498696 | 77.3723528 |
| XXL_VLDL_L | 75.1150047 | 77.1119814 |
| XXL_VLDL_PL | 76.6758003 | 78.2408321 |
| XXL_VLDL_C | 71.5765346 | 73.3322916 |
| XXL_VLDL_CE | 70.1992335 | 71.6435775 |
| XXL_VLDL_FC | 76.979491 | 79.0858324 |
| XXL_VLDL_TG | 76.4113868 | 78.5573453 |
| XL_VLDL_P | 79.6250344 | 81.0204719 |
| XL_VLDL_L | 79.5448983 | 80.9956251 |
| XL_VLDL_PL | 76.7061256 | 77.568133 |
| XL_VLDL_C | 81.4087028 | 83.7791289 |
| XL_VLDL_CE | 83.4283952 | 86.1323091 |
| XL_VLDL_FC | 79.4820862 | 81.4434608 |
| XL_VLDL_TG | 80.2207044 | 81.5567115 |
| L_VLDL_P | 65.7486415 | 66.4649519 |
| L_VLDL_L | 65.9092245 | 66.6492615 |
| L_VLDL_PL | 63.2828785 | 63.7745998 |
| L_VLDL_C | 67.8103357 | 68.9466896 |
| L_VLDL_CE | 65.881665 | 67.1605028 |
| L_VLDL_FC | 70.3224673 | 71.2868938 |
| L_VLDL_TG | 66.255752 | 66.933635 |
| M_VLDL_P | 49.779279 | 50.2494972 |
| M_VLDL_L | 49.1202053 | 49.5923004 |
| M_VLDL_PL | 46.6588475 | 47.0130184 |
| M_VLDL_C | 43.6058508 | 43.9530795 |
| M_VLDL_CE | 39.5890343 | 39.8583961 |
| M_VLDL_FC | 51.1120021 | 51.6378754 |
| M_VLDL_TG | 54.3346828 | 54.9600892 |
| S_VLDL_P | 33.7827011 | 33.6950191 |
| S_VLDL_L | 33.0284012 | 32.89512 |
| S_VLDL_PL | 29.7788245 | 29.5948246 |
| S_VLDL_C | 31.6622888 | 31.4064588 |
| S_VLDL_CE | 33.529671 | 33.2961436 |
| S_VLDL_FC | 31.2261503 | 31.0115549 |
| S_VLDL_TG | 39.9199463 | 40.1761139 |
| XS_VLDL_P | 26.1711385 | 26.0925375 |
| XS_VLDL_L | 26.2745924 | 26.2111408 |
| XS_VLDL_PL | 27.9537422 | 27.7793359 |
| XS_VLDL_C | 27.1858789 | 27.2833624 |
| XS_VLDL_CE | 27.7454348 | 27.895036 |
| XS_VLDL_FC | 26.611448 | 26.5422753 |
| XS_VLDL_TG | 29.6841741 | 29.6340473 |
| IDL_P | 24.4839949 | 24.4364648 |
| IDL_L | 24.7797567 | 24.7516253 |
| IDL_PL | 24.0806001 | 24.1162489 |
| IDL_C | 26.159685 | 26.1531669 |
| IDL_CE | 26.3220057 | 26.3140449 |
| IDL_FC | 26.0320784 | 26.0412505 |
| IDL_TG | 25.2206903 | 25.0290961 |
| L_LDL_P | 25.5379298 | 25.4300924 |
| L_LDL_L | 25.7397183 | 25.6556476 |
| L_LDL_PL | 22.1884754 | 22.2241146 |
| L_LDL_C | 27.9211339 | 27.8186256 |
| L_LDL_CE | 29.3118093 | 29.1490306 |
| L_LDL_FC | 24.4901749 | 24.5377279 |
| L_LDL_TG | 24.4857471 | 24.1882527 |
| M_LDL_P | 26.7076278 | 26.5372388 |
| M_LDL_L | 26.6989124 | 26.5496789 |
| M_LDL_PL | 20.6249593 | 20.5925662 |
| M_LDL_C | 30.109587 | 29.9438435 |
| M_LDL_CE | 33.5762825 | 33.329906 |
| M_LDL_FC | 21.0242553 | 21.0487525 |
| M_LDL_TG | 24.2848467 | 23.7594897 |
| S_LDL_P | 25.3913103 | 25.2195114 |
| S_LDL_L | 25.5035483 | 25.3525773 |
| S_LDL_PL | 18.6322937 | 18.5711357 |
| S_LDL_C | 29.8354792 | 29.6780534 |
| S_LDL_CE | 33.3522874 | 33.1265766 |
| S_LDL_FC | 20.9542286 | 20.9450542 |
| S_LDL_TG | 26.2076689 | 25.6185036 |
| XL_HDL_P | 31.3238982 | 30.8353673 |
| XL_HDL_L | 31.6130266 | 31.1127753 |
| XL_HDL_PL | 32.796077 | 32.2174509 |
| XL_HDL_C | 31.9338368 | 31.5181805 |
| XL_HDL_CE | 30.8321453 | 30.5275509 |
| XL_HDL_FC | 35.4780747 | 34.7519287 |
| XL_HDL_TG | 31.6321724 | 31.8631932 |
| L_HDL_P | 26.8838188 | 26.414529 |
| L_HDL_L | 27.391484 | 26.9085993 |
| L_HDL_PL | 25.2573503 | 24.841228 |
| L_HDL_C | 30.6578085 | 30.0966754 |
| L_HDL_CE | 30.1280582 | 29.6008617 |
| L_HDL_FC | 32.5076923 | 31.8285699 |
| L_HDL_TG | 25.3010461 | 25.1729406 |
| M_HDL_P | 17.8292594 | 18.0117306 |
| M_HDL_L | 18.300813 | 18.4917944 |
| M_HDL_PL | 17.1271273 | 17.1700489 |
| M_HDL_C | 21.4772127 | 21.870333 |
| M_HDL_CE | 21.4161955 | 21.8952173 |
| M_HDL_FC | 22.2455378 | 22.265971 |
| M_HDL_TG | 24.3343142 | 23.5834474 |
| S_HDL_P | 11.7180054 | 12.2228891 |
| S_HDL_L | 11.720344 | 12.2413955 |
| S_HDL_PL | 15.7586902 | 16.193427 |
| S_HDL_C | 12.19465 | 12.8604191 |
| S_HDL_CE | 13.8208863 | 14.4965261 |
| S_HDL_FC | 14.379899 | 14.6868151 |
| S_HDL_TG | 26.8041562 | 26.9110186 |
| VLDL_D | 2.77389552 | 2.77387926 |
| LDL_D | 0.27591447 | 0.27096932 |
| HDL_D | 2.03749424 | 2.00716454 |
| Serum_C | 19.9315104 | 19.9003617 |
| VLDL_C | 34.0272509 | 33.9701097 |
| Remnant_C | 27.7669338 | 27.6868457 |
| LDL_C | 28.892828 | 28.8604257 |
| HDL_C | 18.7727635 | 18.5765655 |
| HDL2_C | 24.0263098 | 23.7726419 |
| HDL3_C | 7.65396033 | 7.57146595 |
| EstC | 19.4788977 | 19.4802237 |
| FreeC | 21.4545866 | 21.3310916 |
| Serum_TG | 36.7550311 | 36.8737536 |
| VLDL_TG | 49.9966331 | 50.4465927 |
| LDL_TG | 24.5764065 | 24.2079206 |
| HDL_TG | 20.7482446 | 20.5181251 |
| TotPG | 16.6066118 | 16.1853022 |
| PC | 18.9674985 | 18.6480753 |
| SM | 16.7622879 | 16.8152572 |
| TotCho | 16.1564815 | 15.9190577 |
| ApoA1 | 11.6807835 | 11.5743579 |
| ApoB | 22.2718842 | 22.1896442 |
| TotFA | 18.381497 | 18.1375654 |
| DHA | 16.4767177 | 16.4317015 |
| LA | 17.3879426 | 17.2744903 |
| FAw3 | 17.8339851 | 17.9868569 |
| FAw6 | 17.4424661 | 17.1711924 |
| PUFA | 17.0075872 | 16.7748706 |
| MUFA | 23.3964174 | 23.0579032 |
| SFA | 18.7364602 | 18.6921158 |
| Lactate | 29.8829816 | 30.9256264 |
| Pyruvate | 24.9622472 | 25.2459315 |
| Citrate | 15.5147851 | 15.3289697 |
| Glycerol | 29.6877535 | 29.6882767 |
| Alanine | 11.5065128 | 11.5823933 |
| Glutamine | 9.87281599 | 9.90516735 |
| Glycine | 15.2336432 | 14.8338588 |
| Histidine | 15.6502765 | 67.1375875 |
| Isoleucine | 21.6321603 | 22.2624303 |
| Leucine | 13.8775598 | 14.2075488 |
| Valine | 15.7300008 | 16.2872408 |
| Phenylaniline | 13.6750929 | 76.3769729 |
| Tyrosine | 14.4351474 | 126.78842 |
| Acetate | 16.8907612 | 32.1749936 |
| bOHBut | 36.40648 | 32.28221 |
| Creatine | 14.95374 | 16.22093 |
| Albumin | 7.667441 | 7.465081 |
| Glycoprotein Acetyls | 11.04558 | 11.21853 |

1. Table showing mean VIPs of clinical covariates in the prediction of GDM status within each ethnicity averaged across 20 model iterations. Standard errors shown in brackets. All model iterations were significant (p value R^2^<0.05, p value Q^2^ < 0.05). Sample sizes for each analysis were as followed: prediction of case status within WEs: 128 cases; prediction of case status within SAs: 286 cases. GDM, Gestational Diabetes Mellitus; MW, Man-Whitney; SA, South Asian; WE, White European.
2. Proportion of outcome variance explained by PLSDA models. Optimised component number selected based upon the significance of pR^2^Y and pQ^2^, the minimisation of RMSEE and the maximisation of R^2^Y. Models included the following covariates: BMI (continuous), age (continuous), multiple pregnancy, parity and smoking status. GDM, Gestational Diabetes Mellitus.
3. Table showing VIP scores from PLSDA models predicting ethnicity in the overall population (n=5339). Model 1: Included covariates of maternal age (years), smoking status, parity, and BMI (continuous). Model 2: Model one covariates and GDM status. Both models were statistically significant (P value R^2^ < 0.05 and P value Q^2^ <0.05). FAw6, N-6 fatty acids; GDM, Gestational Diabetes Mellitus; MUFA, total monounsaturated fatty acids; PUFA, total polyunsaturated fatty acids; SFA, total saturated fatty acids.
